# Proteomics signatures associated with cognitive trajectories: evidence from the English Longitudinal Study of Ageing

**DOI:** 10.64898/2026.03.19.26348791

**Authors:** Jessica Gong, Mikaela Bloomberg, Shaun Scholes, Xinran Hao, Dervis A. Salih, Paola Zaninotto, Andrew Steptoe

**Author notes:** **Corresponding author:** Dr Jessica Gong, Department of Epidemiology and Public Health, University College London, London, UK, 1-19 Torrington Pl, London WC1E 7HB, United Kingdom.

## Abstract

Alzheimer’s disease and related dementias (ADRD) pose a growing global health challenge, with early detection critical to slowing cognitive decline and prevent ADRD. We analyzed high-throughput plasma proteomics in 2,460 cognitively healthy adults from the English Longitudinal Study of Ageing (ELSA) to identify proteins linked to 15-year cognitive trajectories, including verbal fluency, episodic memory, and orientation. Mixed-effect linear models revealed 34 proteins associated with faster orientation decline and 18 with accelerated episodic memory decline. Enrichment analyses implicate extracellular matrix remodeling, immune signaling, apoptosis, and lysosomal-autophagic pathways in cognitive deterioration. Subgroup analyses showed sex-specific effects, highlighting heterogeneity in proteomics signatures in cognitive aging. Notably, ten identified proteins are targets of drugs under clinical investigation, suggesting opportunities for therapeutic repurposing. These findings define a plasma proteomic signature associated with decline in domain-specific cognitive functions, offering promising biomarkers and druggable targets to prevent or slow age-related cognitive decline.

## Introduction

Alzheimer’s disease and related dementias (ADRD) represent major contributors to disability and mortality in older adults.[1, 2] Diagnosis of Alzheimer’s disease (AD) is often preceded by a prolonged preclinical phase characterized by accelerated cognitive decline. As such, preventative research commonly targets cognitive decline with the aim of delaying cognitive impairment and dementia.[3]

The discovery of biomarkers of accelerated cognitive decline could enable early interventions and potentially delay the onset of cognitive impairment and dementia. Moreover, the availability of disease-modifying treatments for ADRD is currently incommensurate with the growing burden of these conditions.[4] Proteomics, the study of proteins and their functions, holds promise for identifying markers and drug targets for ADRD. As proteins drive biological processes, changes in their expression or modification may signal early disease progression,[5] offering insights into molecular changes related to cognitive trajectory.

While several population-based studies have explored proteomic markers for predicting future dementia risk,[6–11] research on plasma proteins in relation to cognitive function has often been constrained by cross-sectional study designs.[12–16] Studies investigating longitudinal associations with cognitive decline have been further limited by small sample sizes,[7, 8, 16–24] case-control study designs,[17] relatively short follow-ups,[18, 23–25] or without excluding individuals with cognitive impairment which could indicate prodromal dementia.[7, 8, 18, 23, 24, 26]

This study leverages high-throughput proteomic data from the English Longitudinal Study of Ageing (ELSA), with repeated cognitive assessments across three cognitive domains (verbal fluency, episodic memory, and orientation) over approximately 15 years, including up to seven biennial measures from 2008 to 2023, to identify the protein signatures associated with cognitive decline among cognitively healthy individuals at baseline, stratified by sex, age, and APOE □4 status. We also investigate the biological pathways and associated drugs currently in clinical testing among identified proteins.

## Results

### Baseline characteristics

Participant selection for the proteomics assays in ELSA is presented in Supplementary Figure 1. At study wave 4 in 2008-09, the 2,460 cognitively normal participants at study baseline included had a mean age of 63.0 years [standard deviation (SD)=8.6], with 56.7% being women, 97.5% of White ethnicity, and 17.8% of the participants carried at least one copy of the APOE ε4 allele (Table 1). The mean raw (i.e., unstandardized) score at wave 4 for verbal fluency, episodic memory, and orientation cognitive domains were 25.8 [SD=5.9], 8.4 [SD=1.4], and 6.0 [SD=0] respectively.

**Table 1.**
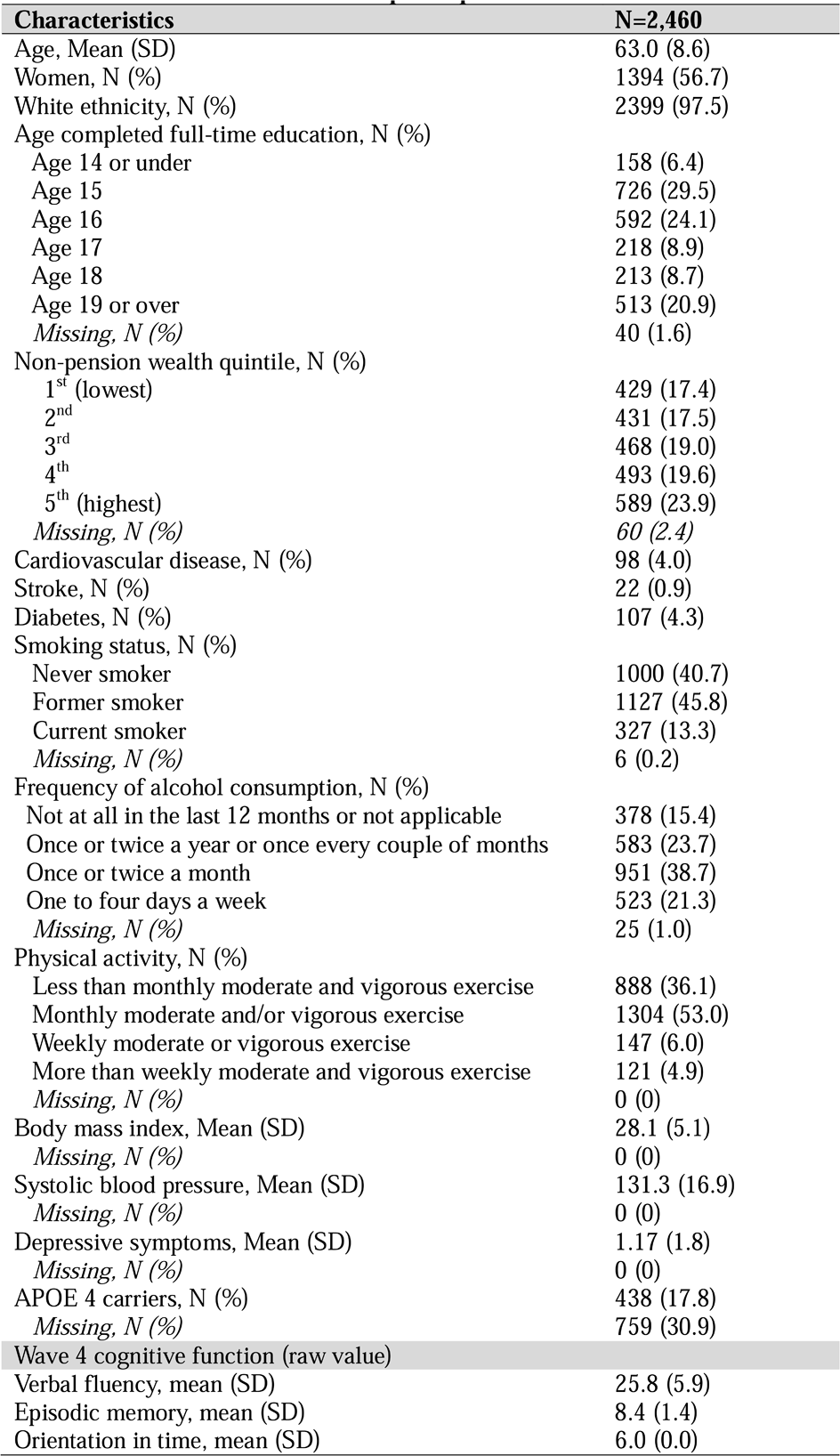
Baseline characteristics of ELSA participants.

### Main analysis: Mixed-effect linear regression for proteins associated with cognitive trajectories

All domains of cognitive function showed decline over age: β (se)=-0.022 (0.001) for verbal fluency, β (se)=-0.028 (0.001) for episodic memory, and β (se)=-0.016 (0.001) for orientation; all p<0.001 (Supplementary Table 1).

We estimated the associations between protein concentration and the trajectory of each cognitive domain. In the fully-adjusted mixed-effect linear regression models (LMMs) (Figure 1), after accounting for false discovery rate (FDR) in multiple testing, the results revealed that higher concentration of 34 proteins (EDA2R, NEFL, BNP, DCN, MMP12, KIM1, MERTK, LAYN, PGF, TNFRSF12A, SCARF2, AMBP, VWC2, TRAIL.R2, GFR.alpha.1 (also known as GFRA1), SCARB2, MMP7, TNFRSF13B, PSG1, RSPO1, VSIG2, TNFRSF11A, SKR3, JAM.B, SFRP1, MSR1, PRELP, SPON2, UNC5C, SMOC2, SLAMF7, REN, TNFRSF10A, GDNFR.alpha.3 (also known as GFRA3)) were associated with steeper decline in orientation. Additionally, higher concentration of 18 proteins (EDA2R, NEFL, KIM1, TNFRSF12A, LAYN, BNP, MMP12, CTSL, DCN, PGF, TMPRSS5, MMP7, SCARF2, MATN3, GFRA3, TF, SKR3, N2DL2) were associated with steeper decline of episodic memory in those cognitively normal at baseline. No protein was associated with decline in verbal fluency.

**Figure 1.**
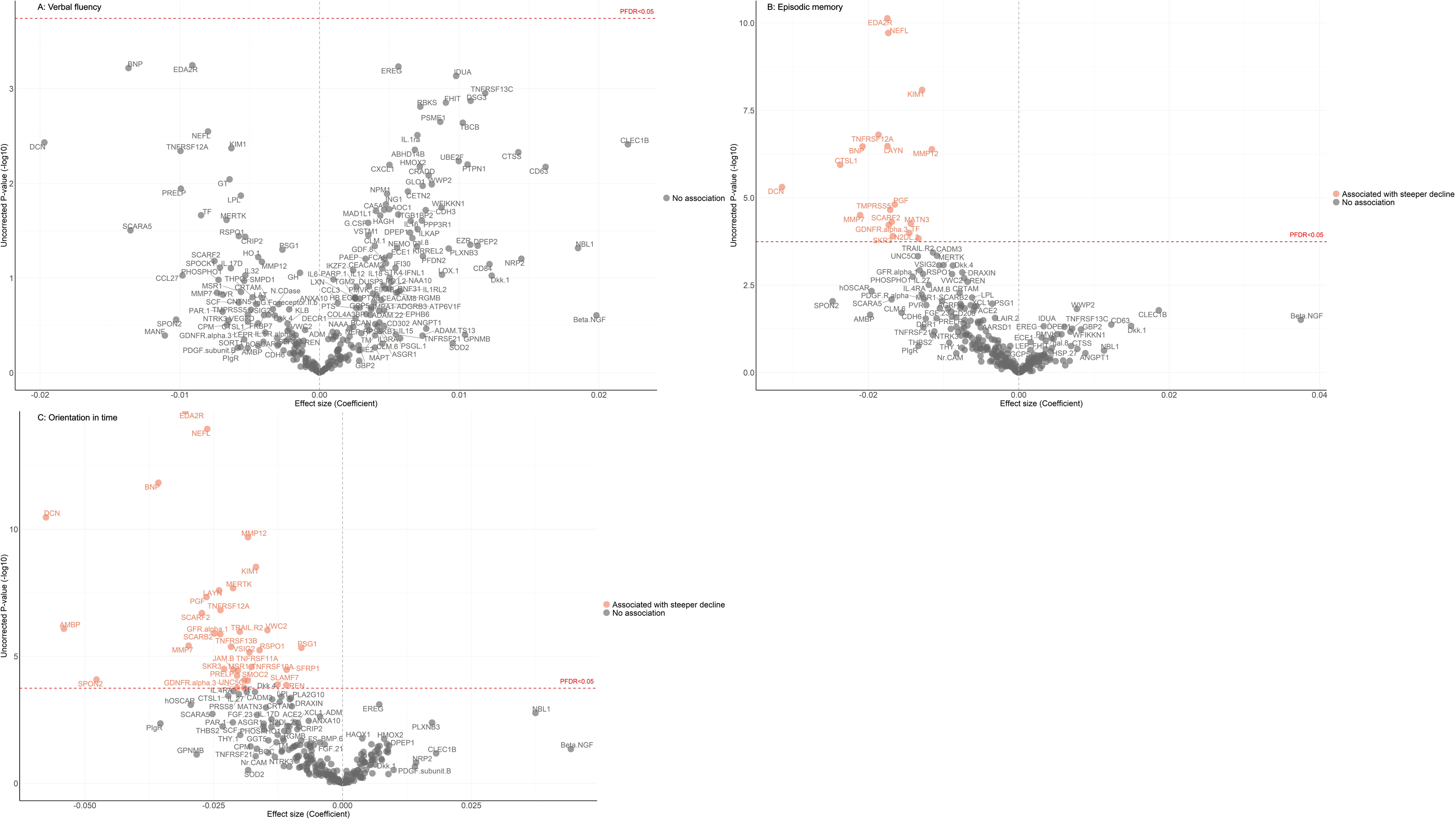
Volcano plots visualizing protein concentration associated with verbal fluency, episodic memory, orientation in time, from fully adjusted mixed-effect linear regression models. Adjusted for baseline age, sex, ethnicity, education, wealth quintile, stroke, diabetes, cardiovascular disease, systolic blood pressure, smoking status, alcohol consumption, physical activity, body mass index, depressive symptoms, allowing for interactions between protein × time since protein measurement.

Minimally adjusted models showed broadly comparable results as the fully adjusted models (Supplementary Figure 2).

### Subgroup analysis: Mixed-effect linear regression by sex, APOE ε4 carriage, and age at baseline

We then conducted a series of subgroup analyses to 1) describe the cognitive trajectories across age; and 2) examined the protein concentration associated with cognitive decline, by sex (women vs men), APOE ε4 carriage (APOE carrier vs non-carrier), and age at baseline (>65 vs ≤65 years of age) (Figure 2).

**Figure 2.**
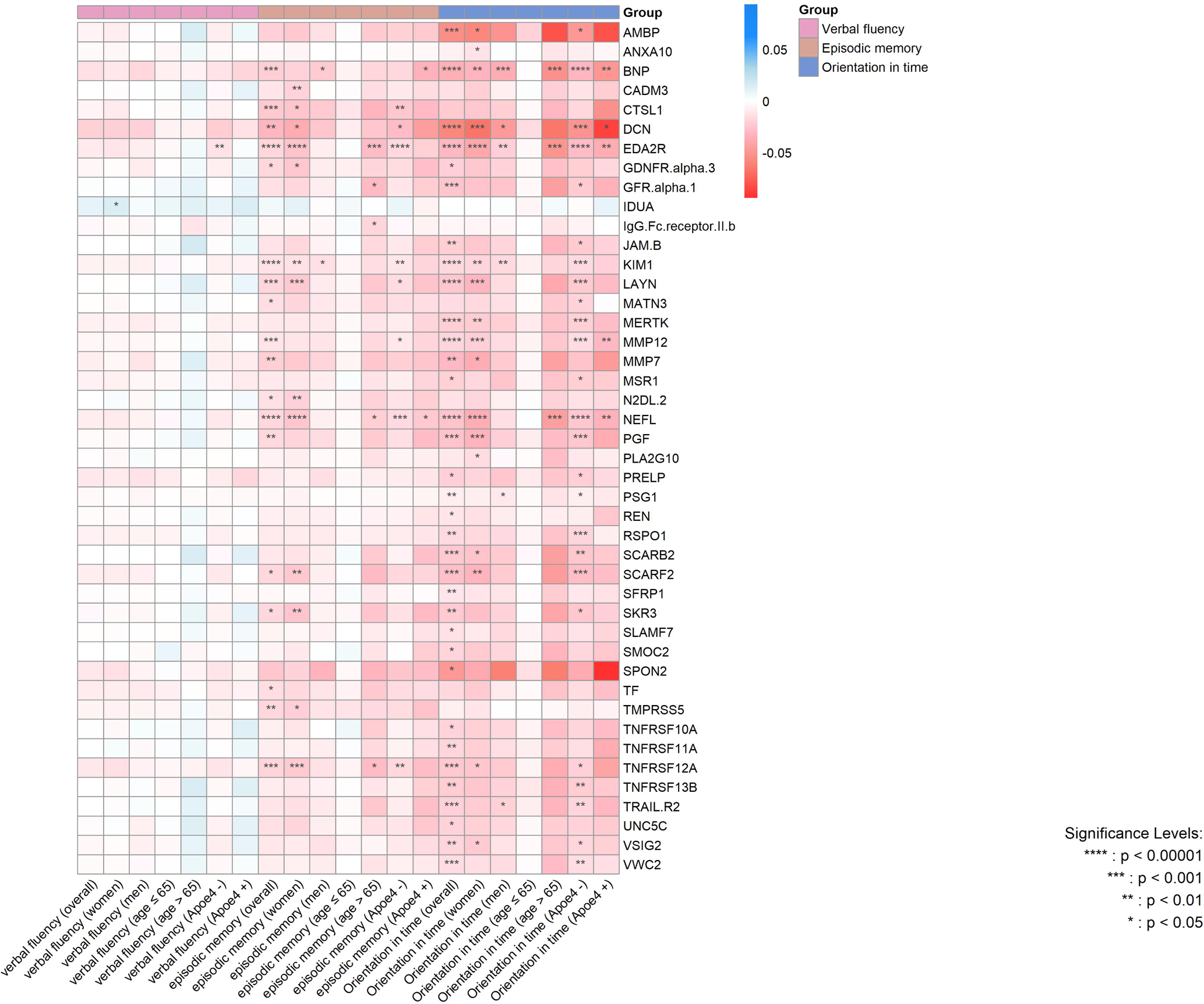
Summary heat map for significant proteins associated with cognitive trajectory across the subgroups, from fully adjusted mixed-effect linear regression models. P values are after false discovery rate (FDR) correction.

There was no sex difference in cognitive decline across age (Supplementary Table 1; Supplementary Figure 3). In the sex-stratified analyses for proteins associated with cognitive trajectories, higher concentration of one protein (KIM1) was identified as being associated with steeper episodic memory decline in women and men; however, higher concentration of 11 proteins (CADM3, CTSL1, DCN, EDA2R, GDNFR.alpha.3, LAYN, N2DL.2, NEFL, SCARF2, SKR3, TMPRSS5, TNFRSF12A) were associated with steeper episodic memory decline in women, and the associations were not significant for men. Conversely, higher concentration of one protein (BNP) was associated with steeper episodic memory decline in men, and not in women. For orientation, higher concentration of four proteins (BNP, DCN, EDA2R, KIM1) were commonly associated with steeper decline in women and men. A total of 12 proteins (AMBP, ANXA10, LAYN, MERTK, MMP12, MMP7, NEFL, PGF, PLA2G10, SCARB2, SCARF2, TNFRSF12A, VSIG2), where higher concentration of these proteins were associated with steeper orientation decline in women but not in men, and two proteins (PSG1, TRAIL.R2) were associated with steeper orientation decline in men but not in women. There was no difference in cognitive decline across all domains between APOE ε4 and non-carriers over age (Supplementary Table 1; Supplementary Figure 4). Protein-association analyses stratified by APOE ε4 carriage revealed that one protein (NEFL) was associated with steeper decline in episodic memory in both carriers and non-carriers. Among individuals with no APOE ε4 allele, higher concentrations of seven proteins (CTSL1, DCN, EDA2R, KIM1, LAYN, MMP12, TNFRSF12A) were associated with steeper decline in episodic memory, but these associations were not observed for carriers. Conversely BNP was associated with steeper decline in episodic memory in APOE ε4 carriers but not observed in non-carriers. For decline in orientation, higher concentration of five proteins (BNP, DCN, EDA2R, MMP12, NEFL) were associated with steeper decline for APOE ε4 carriers and non-carriers. A total of 21 proteins (AMBP, GFR.alpha.1, JAM.B, KIM1, LAYN, MATN3, MERTK, MMP12, MSR1, PGF, PRELP, PSG1, RSPO1, SCARB2, SCARF2, SKR3, TNFRSF12A, TNFRSF13B, TRAIL.R2, VSIG2, VWC2) showed that higher concentrations were associated with steeper decline in orientation among non-carriers but not observed in carriers.

In the stratified analysis for protein associated with cognitive trajectory by age group at baseline, significant associations were only observed among those greater than 65 years of age: higher concentrations of EDA2R, GFR.alpha.1, IgG.Fc.receptor.II.b, NEFL, TNFRSF12A were associated with steeper decline in episodic memory; and higher concentrations of BNP, EDA2R, NEFL were associated with steeper decline in orientation.

### Enrichment network analysis

Subsequently, we conducted enrichment network analysis to further understand the biological mechanisms underlying the signatures of the proteins identified from the main and subgroup analyses separately by cognitive domains, using EnrichR-KG, and confirmed by gprofiler2 and Ingenuity Pathway Analysis (IPA, Qiagen).

The network analysis of proteins associated with episodic memory reveals key functional connections centered on extracellular matrix (ECM) remodeling, proteolysis, and wound healing (Figure 3). CTSL1, MMP7, and MMP12 are consistently involved in processes such as cellular component disassembly, ECM disassembly, and organization, underscoring their roles in tissue remodeling. DCN and MATN3 further contribute to ECM organization and proteoglycan-related processes, while ACVRL1 and MMP12 are linked to wound healing, cell migration, and the negative regulation of cell-matrix adhesion. Pathway analysis supports these roles, with ACVRL1 involved in cytokine interactions and DCN and CTSL1 associated with proteoglycans in cancer, highlighting potential functions in immune signaling and tumor progression. Reactome pathways emphasize the involvement of these proteins in ECM dynamics, including collagen degradation and matrix remodeling, with CTSL1 and MMP7 also playing roles in collagen formation and fibril assembly. Notably, CTSL1 is linked to pathways related to SARS-CoV-2 infection, suggesting a broader relevance in viral entry and disease mechanisms. This subnetwork illustrates a tightly coordinated system regulating ECM integrity, tissue repair, and proteolytic processes, with the repeated involvement of MMPs and CTSL1 in matrix degradation suggesting their potential as therapeutic targets for conditions involving excessive tissue remodeling.

**Figure 3.**
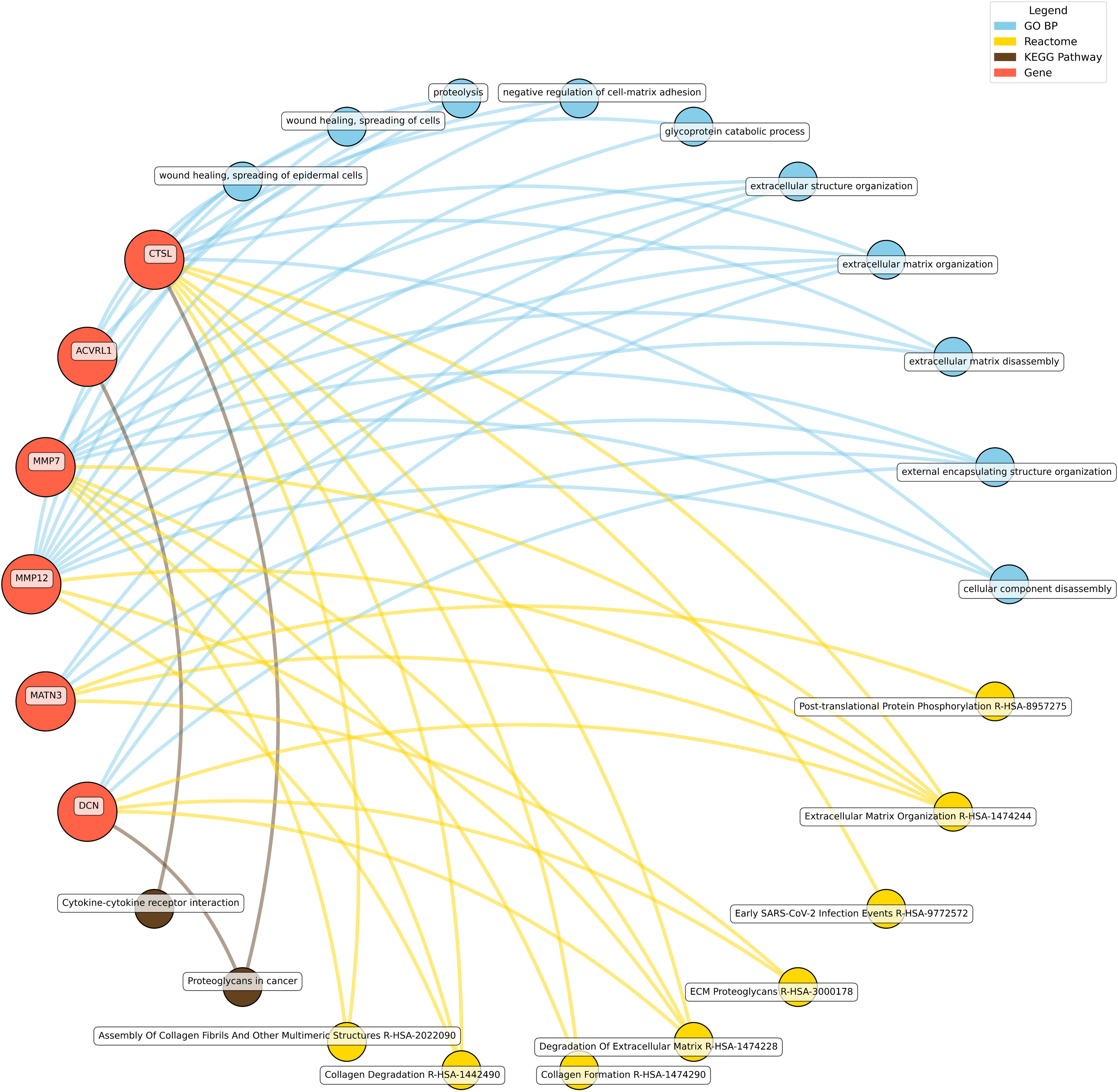
Network analysis for pathways enriched in significant proteins for episodic memory domain from the fully adjusted mixed-effect linear models. GO BP gene ontology – biological processes, Reactome a database of reactions, pathways and biological processes, KEGG Kyoto Encyclopedia of genes and genomes. The ‘node’ indicates the gene name corresponding to the protein, or the biological term extracted from each bioinformatics library, the ‘edge’ connects protein to their enriched term.

The enrichment network of proteins associated with cognitive decline in orientation identifies two key functional themes: apoptosis regulation via TNF receptor signaling and ECM organization/remodeling (Figure 4; Supplementary Figures 5-6). These processes are interconnected through genes involved in both cell death pathways and tissue maintenance. TNF receptor superfamily genes play central roles in apoptotic signaling, particularly in TRAIL-activated apoptosis and extrinsic apoptotic signaling and pro-inflammatory signaling (Supplementary Figures 6-8). Pathway analysis links these genes to necroptosis, NF-κB signaling, cytokine interactions, and viral infection responses (e.g., Influenza A), with Reactome pathways highlighting their roles in caspase activation, TRAIL signaling, and p53-mediated regulation of death receptors. A distinct set of proteins (MMP7, MMP12, DCN, SMOC2, and JAM2) was found to be primarily involved in ECM organization, wound healing, and external structure formation. SFRP1, DCN, and SMOC2 are implicated in angiogenesis regulation, while MMP7 and SFRP1 interact with the Wnt signaling pathway, linking them to developmental and tissue repair processes. Reactome pathways reinforce the involvement of these genes in ECM degradation, crucial for tissue remodeling and fibrosis (Supplementary Figure 5). Notably, TNFRSF10A and TRAIL-R2 bridge apoptosis and ECM regulation, influencing cell death and vascular interactions. MMPs also participate in apoptotic signaling, highlighting their dual role in ECM remodeling and cell death modulation. The network further reveals the involvement of TNF receptor genes in viral infection pathways and MMPs in Wnt signaling, suggesting broader implications for host-pathogen interactions, arthritis and cancer progression (Supplementary Figure 6). The integration of apoptosis, NF-κB activation, and ECM remodeling underscores the relevance of this subnetwork to cancer, inflammation, and tissue regeneration. Given their roles in immune modulation and cell death, the TNF receptor family genes are promising targets for cancer immunotherapy and treatment of autoimmune disorders.

**Figure 4.**
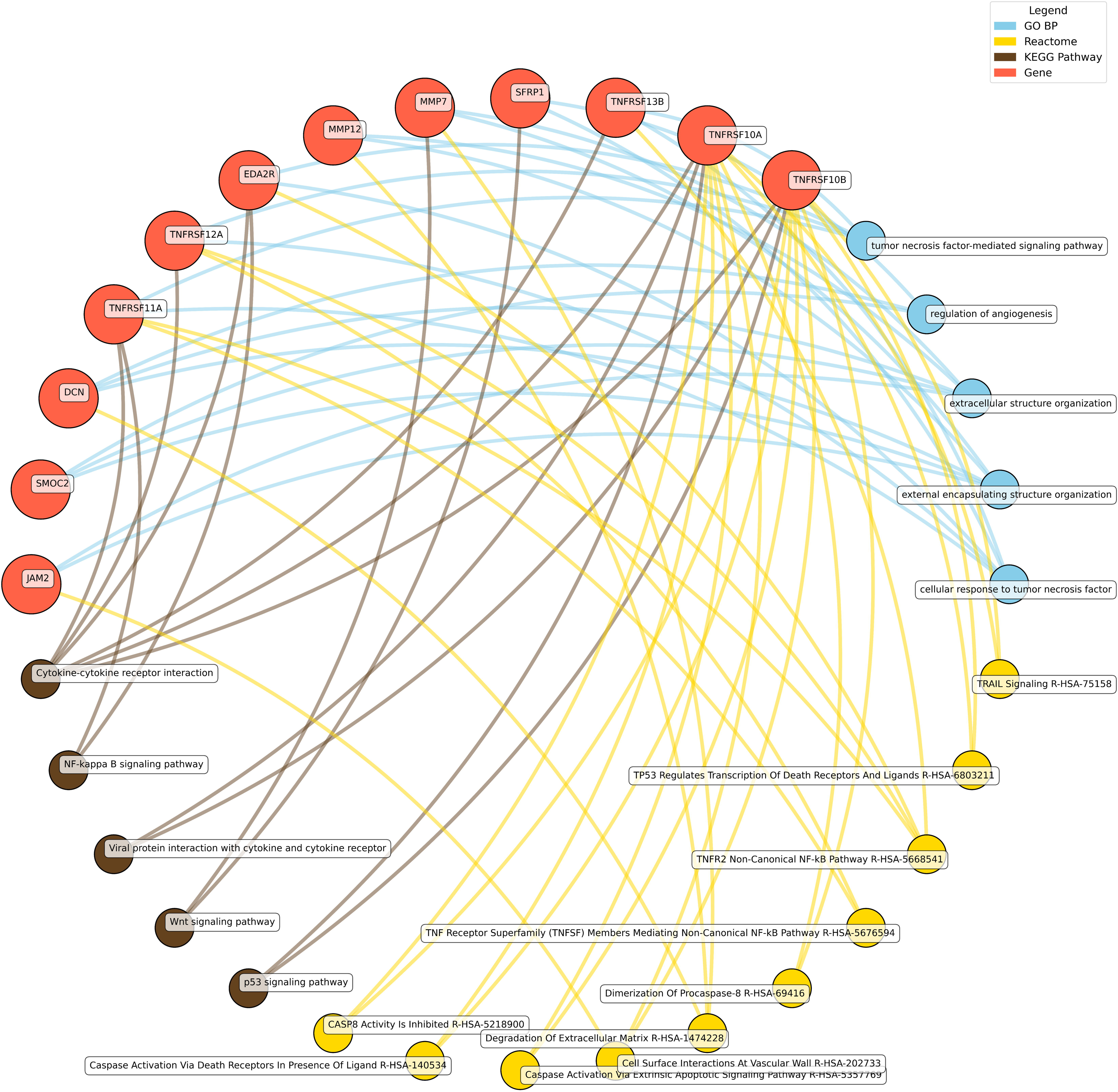
Network analysis for pathways enriched in significant proteins for orientation domain from the fully adjusted mixed-effect linear models. GO BP gene ontology – biological processes, Reactome a database of reactions, pathways and biological processes, KEGG Kyoto Encyclopedia of genes and genomes. The ‘node’ indicates the gene name corresponding to the protein, or the biological term extracted from each bioinformatics library, the ‘edge’ connects protein to their enriched term.

To gain further insight into the mechanisms underlying the change in these proteins associated with cognitive decline, we performed an analysis of upstream regulators of the proteins indicates this network is coordinated by factors such as the pro-inflammatory cytokine IL-6, anti-inflammatory cytokine IL-10, immunomodulator TNFSF13B and the Parkinson’s disease familial and risk factor LRRK2 (Supplementary Figure 8). We see gene variants for *LPL, SCARB2, MSR1, LAYN* and *PPP3R1* associated with blood lipids, diabetes, metabolites, Parkinson’s disease and AD from GWAS associations. *LPL* is colocalized with an Expression Quantitative Trait Loci (eQTL) in the blood and nerves, *SCARB2* is colocalized with an eQTL in fibroblasts and spinal cord, *MSR1* in the blood, *LAYN* in the putamen, and *PPP3R1* in the arteries, fibroblasts and myeloid cells, from mining data from the Genotype-Tissue Expression (GTEx). With cell-type specificity in the brain, we see that five proteins are part of ageing related glial transcriptome networks in mice or humans: LPL, SCARB2, MSR1, LAYN and PPP3R1 (expressed by microglia or oligodendrocytes from bulk or single-cell RNA-seq. An independent dataset from the Religious Order Study and the Rush Memory and Aging Project (ROSMAP) confirms expression of these 5 proteins in microglia and/or oligodendrocytes (Supplementary Figure 9). Finally, *SCARB2* is colocalized with an eQTL acting in oligodendrocytes, and *MSR1* in microglia (Supplementary Figure 10). Thus, eQTLs associated with these genes may contribute to nervous system function via neurons, the vasculature, immune system and oligodendrocytes.

### Open target for drug identification

Using the full list of proteins identified in the main and subgroup analyses, by querying the Open Targets Platform, we found that 10 proteins (MMP7, MMP12, PGF, SKR3 (also known as ACVRL1), SLAMF7, REN, MERTK, GFRA1, TNFRSF10A, and TRAIL-R2) currently have drugs in clinical testing (Figure 5). These 10 proteins are linked to a total of 25 drugs across 532 registered clinical trials.

**Figure 5.**
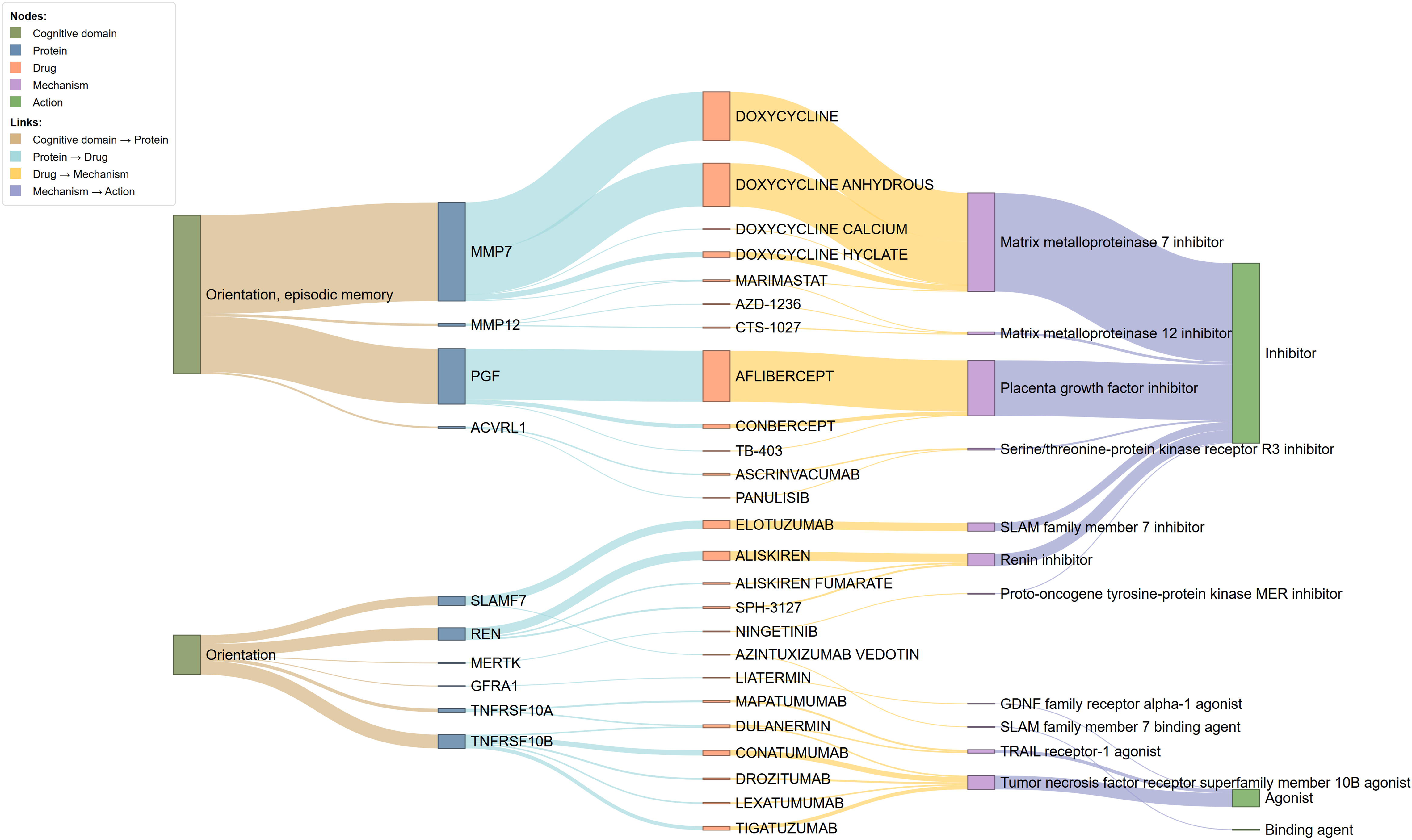
Sankey diagram showing existing drugs linked to the identified proteins associated with cognitive decline by domain, drug name, mechanism of action, and action type.

Higher concentrations of six proteins (SLAMF7, REN, MERTK, GFRA1, TNFRSF10A, TRAIL-R2) correlated with steeper declines in orientation alone, while elevated levels of four proteins (MMP7, MMP12, PGF, SKR3) were associated with accelerated declines in both orientation and episodic memory.

Among the 25 drugs identified, three drugs in nine trials targeting two proteins (MMP7, GFRA1) are specifically targeting AD, familial amyloid neuropathy, or Parkinson’s disease. The remaining drugs target diverse conditions, including various cancers, kidney disease, infectious diseases, pulmonary diseases, ocular diseases, and cardiometabolic disorders. These therapies encompass small molecules, antibodies, antibody-drug conjugates, and proteins, with and nearly a third (30.8%, 164/532) are in Phase III or IV trials (Supplementary Table 2).

Mechanistically, 17 drugs targeting seven proteins function as inhibitors, suppressing the activity or expression of their target proteins. The remaining drugs act as binding agents or agonists, enhancing the activity or expression of their respective targets.

## Discussion

### Summary of key findings

To our knowledge, this study is among the largest to date, featuring 15 years of follow-up to investigate proteome-wide associations with longitudinal cognitive decline across three cognitive domains in individuals who were cognitively healthy at baseline, within a nationally representative population-based cohort. In the primary analysis, elevated levels of 34 proteins were associated with declines in orientation, while 18 proteins were linked to declines in episodic memory. These proteins reflected both overlapping and distinct biological pathways, with ten already targeted by drugs currently undergoing clinical testing. Subgroup analyses further revealed sex-specific effects and patterns dependent on APOE □4 status, underscoring the heterogeneity of proteomic signatures in cognitive aging.

### Shared biological pathway between orientation and episodic memory decline

#### 1. Neuroinflammation and immune dysregulation

Several proteins associated with cognitive decline are found to be implicated in neuroinflammation and immune dysregulation. TNFRSF12A, a member of the tumor necrosis factor receptor superfamily, promotes pro-inflammatory signaling and has been linked to synaptic loss in AD.[27] Similarly, TNFRSF11A, TNFRSF10A are key regulators of innate immunity and inflammatory signaling implicated in autoinflammatory disorders,[28, 29] and may play a role in modulating hippocampal and prefrontal circuits in chronic inflammation. MERTK, a phagocytic receptor on microglia, is implicated in amyloid-β clearance deficits in AD.[30, 31]

Additionally, KIM1 (also known as HAVCR1), though typically a renal marker increased in the proximal tubules of damaged or diseased kidneys,[32] also plays a key functional role in maintaining regulatory B-cell homeostasis, including their generation, expansion, and immunosuppressive activity,[33] and may be upregulated in brain injury and may exacerbate neuroinflammatory and autoimmune responses.[34] With the enrichment analysis highlighting several enriched biological immune and inflammatory pathways such as cytokine-cytokine receptor interactions, these findings align with evidence that persistent inflammation disrupts neuropathic and neurodegenerative mechanisms.[35, 36]

#### 2. Axonal injury and synaptic dysfunction

Our findings on NEFL align with established literature, reinforcing their roles as critical protein markers in neurodegeneration and cognitive decline. NEFL serves as a robust non-specific biomarker for neuronal damage and axonal injury,[37–39] and is a marker of neurodegeneration across a variety of conditions,[40] including ADRD,[40–42] multiple sclerosis,[40, 43] and other neurodegenerative disorders.[40]

EDA2R is involved in apoptosis, may further exacerbate neuronal loss, and have been associated with poorer cognitive ability partially mediated by smaller total brain volume.[13] This parallels studies showing that axonal degeneration precedes overt cognitive symptoms in preclinical AD.[44]

SLAMF7, a synaptic glycoprotein, modulates neuronal-glial communication, has been shown to implicate multiple sclerosis[45, 46] and APOE-related alteration.[47] GFRA3, functions as a co-receptor that partners with the RET tyrosine kinase to mediate signaling by glial cell line-derived neurotrophic factor (GDNF).[33, 48] This complex is critical for the development, maintenance, and survival of sensory and sympathetic neurons in the peripheral nervous system.[33, 48]

UNC5C, a dependence receptor for netrin-1, partners with the Deleted in Colorectal Cancer (DCC) receptor to mediate repulsive axon guidance. Upon netrin binding, the UNC5C-DCC complex triggers cytoskeletal remodeling in neuronal growth cones, steering axons away from inhibitory cues during neural circuit formation in the developing nervous system.[33, 49] These proteins collectively underscore the role of axonal integrity in maintaining cognitive function.

#### 3. Extracellular matrix integrity, blood-brain barrier and vascular dysfunction

Proteins such as MMP12, MMP7 (both matrix metalloproteinases), and DCN regulate ECM integrity and are involved in Blood-Brain Barrier (BBB) dysfunction. MMPs degrade BBB tight junction proteins, are upregulated in cerebral small vessel disease,[50] contributing to BBB leakage and neurovascular dysfunction, a hallmark of vascular contributions to cognitive impairment.[6, 51] Consistently, we found MMP12 associated with greater risk of vascular dementia in our previous study.[6]

PGF (placental growth factor), a pro-angiogenic factor, may amplify BBB disruption, and accelerate amyloid-β deposition, and tau pathology,[52] linking vascular and neurodegenerative pathways. Further, elevated levels of BNP and REN, established biomarkers of cardiovascular stress and hypertensive encephalopathy, are correlated with microvascular damage and cerebral hypoperfusion, processes that exacerbate cognitive decline.[53–56]

Meanwhile, MATN3 modulates ECM-cell interactions,[57] potentially influencing synaptic plasticity. Further, DCN and PRELP also regulate ECM stability,[58, 59] and SPON2[60] and SMOC2[61] modulate ECM-cell adhesion. JAMB is a junctional adhesion protein that mediates heterotypic cell-cell interactions.[33, 62]

Dysregulation of ECM proteins is increasingly recognized as a driver of memory decline in both AD and vascular dementia.[63] BBB degeneration emerges as a pivotal mechanism directly linking protein dysregulation to accelerated cognitive decline, underscoring its central role in the interplay between vascular dysfunction and neurodegeneration.

### Distinct mechanisms underlying orientation decline: broader brain network disconnect

Proteins such as TRAIL-R2 (also known as TNFRSF10B), RSPO1, SFRP1, and JAM-B are uniquely associated with orientation in time decline through distinct yet interconnected mechanisms that impact neural integrity and function. TRAIL-R2, a member of the death receptor family, induces apoptosis primarily through caspase activation. In the central nervous system, aberrant activation of TRAIL-R2 has been associated with neuronal loss in regions critical for cognitive processing.[64] Notably, a study demonstrated that aberrant TRAIL-R2 signaling contributes to neurodegeneration, supporting its role in the pathogenesis of AD.[65]

In a related pathway, RSPO1 and SFRP1, modulators of the Wnt signaling cascade, impact synaptic plasticity and neurogenesis. Dysregulated Wnt signaling may contribute to synaptic dysfunction and degeneration, thereby accelerating the progression of AD.[66] As highlighted in previous literature, dysregulation of this pathway can contribute to broader cognitive deficits.[67]

Moreover, JAM-B, a key component of the BBB, plays an essential role in maintaining neurovascular integrity.[68] Breakdown of the BBB can lead to neuroinflammation and neurovascular uncoupling,[69] processes that significantly compromise cognitive functions in the parietal and frontal cortices,[70, 71] regions responsible for attention and executive function.[72, 73]

Collectively, these findings underscore the crucial roles of apoptosis, synaptic dysfunction, and BBB integrity in maintaining orientation in time, and their disruption can lead to significant cognitive impairment. Notably, impairment in orientation often signals more severe cognitive decline compared to deficits in other cognitive domains.[74] The presence of a larger number of associated proteins suggests a wider array of underlying biological processes contributing to progressive cognitive deterioration. As such, this process may represent an early and more pervasive pathological mechanism that accelerates cognitive decline.

### Proteins uniquely associated with episodic memory decline: possible hippocampal-centric mechanisms

Emerging evidence implicates lysosomal dysfunction and impaired autophagy as central to hippocampal neurodegeneration and episodic memory decline. The lysosomal protease CTSL1,[33] essential for breaking down amyloid-β and hyperphosphorylated tau, is impaired in AD, directly linking lysosomal failure to AD pathogenesis.[75]

In parallel, TMPRSS5, a neural transmembrane serine protease, has been associated with proteolytic dysregulation,[76] and has been identified in central nervous system,[76] its mechanistic role in neurodegeneration remains poorly elucidated, necessitating deeper investigation into its cellular functions and molecular pathways. Several previous studies also highlighted the potential role of autophagy in neurodegeneration.[6, 8]

Iron homeostasis further intersects with hippocampal dysfunction. Transferrin (TF), critical for maintaining iron balance, supports synaptic plasticity and neurotransmitter synthesis,[77, 78] is implicated in function of hippocampus.[79] Dysregulated TF leads to iron accumulation and oxidative stress, impairing memory-related signaling.[80] A study found that excess hippocampal iron suppresses furin and reduces BDNF maturation,[80] while another showed that iron deficiency also compromises spatial memory in mouse hippocampus.[81]

Taken together, these proteins highlight converging mechanisms, lysosomal-autophagic deficits, iron dysregulation, and impaired proteolytic remodeling, that undermine hippocampal resilience that may act via microglia, oligodendrocytes and the vasculature. By promoting toxic protein accumulation and contributing to disruption of synaptic integrity, they contribute to episodic memory loss and reveal molecular targets for interventions aimed at restoring proteostasis, iron balance, and lysosomal function.

### Sex disparities in protein-cognitive decline associations

Regarding sex differences, higher concentrations of several proteins were associated with steeper declines in episodic memory and orientation among women, highlighting potential sex-specific proteomic signatures underlying cognitive decline. The role of estrogen may be crucial in these pathways, as it modulates TNF receptor activity and suppresses pro-inflammatory cytokines.[82, 83] Women may benefit from the neuroprotective effects of estrogen acting synergistically with TNF pathways, exhibiting slower memory decline before menopause. However, after menopause, estrogen levels drop sharply, potentially exacerbating neuroinflammation and oxidative stress and altering the expression of proteins such as MMPs, TNF receptors, and growth factors involved in neuronal maintenance and immune signaling.[84] Furthermore, women generally have a more reactive immune system, which may increase their susceptibility to chronic low-grade inflammation and age-related immune dysregulation, thereby impacting neurodegeneration.[85] Dysfunction of this pathway may be more pronounced in women, leading to accumulation of cellular debris and increased neuroinflammation. Additionally, some immune-related genes are X-linked, potentially leading to sex-biased protein expression.[86] The female-specific benefit likely arises from estrogen’s interactions with these pathways, combined with X-chromosome-linked gene expression in females, which together enhance neuroprotection. Future research should prioritize the inclusion of directly measured sex hormone levels and proxies for hormonal therapy exposure to clarify their mechanistic interactions with these protein pathways and to advance sex-specific therapeutic strategies aimed at neuroprotection in cognitive aging.

### Drugs targeting identified proteins

Several therapeutics under clinical investigation specifically target proteins identified in this study. Although originally developed for cancer, inflammation, infection, or cardiometabolic diseases, these agents act on pathways linked to neurodegeneration, highlighting growing interest in drug repurposing for dementia and related neurocognitive disorders.[87]

For example, matrix metalloproteinase inhibitors such as marimastat and AZD-1236, designed to inhibit MMP7 and MMP12 respectively, may mitigate blood-brain barrier breakdown and neuroinflammation,[88] processes implicated in both episodic memory and orientation cognitive decline.

Similarly, aflibercept, a VEGF pathway inhibitor targeting placental growth factor (PGF), has been used in ophthalmic and oncologic settings but may hold potential for reducing pathological angiogenesis and neurovascular dysfunction contributing to cognitive impairment.[89]

Drugs modulating immune pathways, such as elotuzumab (a SLAMF7 inhibitor) may influence neuroimmune activation.[90] Additionally, agents like lexatumumab and tigatuzumab, targeting TRAIL receptor pathways, offer intriguing links to apoptosis regulation in neural tissues.[91, 92] While these therapies were not originally developed for neurodegenerative diseases, their mechanisms align with pathways implicated in hippocampal dysfunction, synaptic loss, and neurovascular compromise.

### Evidence from previous research on proteomics and cognitive functions

Previous research on the associations between proteomic signatures and cognitive decline over time has been limited. A recent study from the UK Biobank using Olink proteomics assays reported cross-sectional associations between proteins and several cognitive functions including fluid intelligence, reaction time, prospective memory.[12] The authors concluded that GDF15 and CDCP1 exhibited the most significant associations with cognitive function traits.[12] In our previous study investigating proteomic signatures of dementia risk, we identified several proteins associated with greater risk of dementia.[6] Notably, four of these proteins (NEFL, EDA2R, KIM1, and MMP12) also demonstrated significant associations with cognitive decline in the current analysis, underscoring their potential role as shared biomarkers across dementia-related phenotypes.

In the Whitehall II study,[7] proteomics data derived from the SomaScan platform were used to assess the associations with cognitive decline across four domains: executive function, memory, and phonemic and semantic fluency. The analysis identified several proteins linked to immune system activation, BBB dysfunction, vascular pathology, and central insulin resistance. A separate validation analysis from Whitehall II to validate proteins associated with dementia from the Atherosclerosis Risk in Communities (ARIC) study highlighted four specific proteins - DNAJB9, GDF15, HSPA1B, and MMP19 - as being associated with cognitive decline,[8] though these associations was attenuated after adjusting for multiple comparison. This subset of proteins, which play roles in immune function, and ECM organization, may reflect peripheral biological processes underlying early cognitive changes that precede dementia.[8]

### Strengths and limitations

This study has several notable strengths. First, it benefits from a large sample size of cognitively healthy individuals with approximately 15 years of follow-up in a large-scale nationally representative cohort, with the inclusion of numerous highly relevant proteins for studying neurological processes using three Olink Target 96 panels. Second, this study includes up to seven repeated administrations of well-validated cognitive assessments, more than most prior research in this area. Repeated assessments of cognitive function are crucial, as they offer a nuanced understanding of long-term cognitive trajectories, which generally change subtly over extended periods and may provide further understanding into potential causal relationships. Additionally, we modelled cognitive domains separately as each measure may have different sensitivity to change,[93] and these cognitive domains measure different cognitive abilities and may contribute to different aspects of disease etiology.[94, 95] Lastly, the agnostic approach to identifying proteins linked to cognitive decline generates new hypotheses about biological mechanisms, which can then be tested in experimental models and integrated into the drug discovery pipeline.

However, the study also has limitations that should be acknowledged. First, we lacked a suitable validation cohort, as no other studies have assessed cognitive function in a comparable manner or utilized similar proteomics panels. Second, while the included proteins are highly relevant to cognitive decline, they represent only a small fraction of the entire known human proteome,[96] and therefore, other relevant proteins for cognitive decline may have been missed. Further, post-translational modifications (e.g., phosphorylation, glycosylation) and protein isoforms can significantly influence protein function, but these modifications are often not captured in standard proteomic assays. The inclusion of protein modifications and isoforms in future studies could help identify more specific biomarkers and functional insights into how these proteins contribute to cognitive decline. Third, although the longitudinal nature of the study allows for better insight into temporal relationships, it still cannot establish causality definitively. There may be reverse causality (e.g., cognitive decline influencing protein levels), or unmeasured variables may be driving both cognitive decline and proteomic alterations. Finally, while well-validated, the tests of cognitive function used in ELSA were brief and based on isolated tasks,[97] and having multiple tests per domain is preferable.[98]

## Conclusion

In conclusion, this study addressed key gaps by providing robust, longitudinal evidence of proteomic signatures associated with cognitive decline trajectories among those cognitively healthy at study baseline over a 15-year period, with a focus on the underlying biological mechanisms revealed through enrichment network analysis. The identified proteins, particularly those involved in ECM remodeling, proteolysis, neurovascular changes and immune modulation, highlight key functional connections that contribute to both the progression of cognitive decline and the regulation of neuroinflammation. Importantly, many of these proteins play crucial roles in balancing immune responses in microglia, oligodendrocytes and the vascular system, offering new insights into the molecular underpinnings of cognitive decline. Importantly, several proteins identified in this study are already targeted by drugs currently under clinical investigation for other conditions, offering immediate potential for therapeutic repurposing. Further investigation may offer critical insights into the molecular mechanisms underlying cognitive decline and aid in identifying early biomarkers for diagnosis and therapeutic intervention to prevent and slow the progression of cognitive decline.

## Online methods

### Study cohort

The study utilized data from the English Longitudinal Study of Ageing (ELSA), a nationally representative longitudinal study of individuals aged 50 and above, along with their partners, living in England. ELSA began its first wave of data collection in 2002-03 (wave 1), using a combination of in-person interviews and self-reported paper questionnaires.[99] Data collection continued biennially with the most recent wave of data collected in 2021-23 (wave 10). Blood sample collection was introduced in 2004 during the nurse visit (wave 2), with subsequent collections occurring every four years. The present study utilizes blood samples collected during the wave 4 nurse visit (2008-09), which constitutes the baseline wave of this study, and cognitive data from cognitive testing administered at waves 4-10 (2008-09 to 2021-23).

### Proteomics data

For the proteomics assays, three Olink™ Target 96 panels were selected: Cardiovascular II (CVDII), Neurology I (NEUI), and Neurology Exploratory (NEX), which together cover a total of 276 distinct proteins (Supplementary Table 3). The frozen samples were sent to Olink for aliquoting, plating, and assay execution. These assays include built-in quality control measures, with four internal controls added to each sample, along with additional external controls. Protein concentrations were measured using Olink’s standardized Normalized Protein eXpression (NPX) values, which are reported on a Log_2_ scale. The quality control procedures for the proteomics data have been previously described.[6]

For the proteomic analysis in ELSA, blood samples collected during the wave 4 nurse visit in 2008-09 were used (considered the study baseline here). Participants who either died within two years of the wave 4 nurse visit (N=134) or were lost to follow-up (missing at least two consecutive waves) (N=1,340) were excluded. A total of 3,325 frozen plasma blood samples at -80°C were sent to Olink for aliquoting, plating, and proteomics analysis. A total of 3,305 samples from wave 4 were viable for proteomic profiling. The final analysis included a combined dataset across three Olink panels of 3,262 participants who underwent proteomic assays after stringent sample quality control.

### Baseline cognitive status

Baseline all-cause dementia was determined using a dementia algorithm developed for the ELSA study, which integrates data from self-reported physician diagnoses, informant-reported cognitive assessments using the Informant Questionnaire on Cognitive Decline in the Elderly (IQCODE), as well as hospital admission and mortality records. More details on the dementia algorithm can be found elsewhere.[6] Individuals were considered to have cognitive impairment at baseline if they scored 1.5 standard deviations (SD) or more below the mean in at least one of the cognitive domains at protein assessment in wave 4, criteria that are commonly applied for identification of cognitive impairment when clinical assessment is not possible.[100] Subsequently, baseline cognitive status was determined using a combination of algorithmically derived all-cause dementia and the objective cognitive assessments described above. Baseline cognitive status was categorized as cognitively normal or cognitively impaired, with cognitively impaired defined as having dementia at study baseline (according to the dementia algorithm) and/or a cognitive score at baseline that was 1.5 SD or more below the mean at wave 4. Those deemed cognitively impaired were excluded from the analyses (N=802), yielding a final analytical sample size of 2,460 cognitively healthy individuals.

### Assessments of cognitive function

Verbal fluency was evaluated using an animal naming task, where participants were asked to name as many animals as possible within one minute.

Episodic memory was assessed in each interview using the Consortium to Establish a Registry for Alzheimer’s Disease (CERAD) immediate and delayed recall tasks. Participants were orally presented with a list of ten words and were asked to recall as many as possible immediately after hearing the list and again after a delay of approximately five minutes, during which they answered other survey questions. Scores from both recall tasks were summed (range 0-20).

Orientation in time was measured by asking participants to report the current date (day, month, and year) and the day of the week, with responses summed up to produce a total orientation score.

Each of these cognitive tests was administered in every wave of ELSA, except for the animal naming task used to assess verbal fluency, which was not administered in wave 6 (2012-13). The three cognitive measures were standardized within the study sample to have a mean of 0 and a standard deviation of 1.

### Baseline indicators

Sociodemographic factors included chronological age (calculated from the difference between wave 4 nurse visit and participant’s date of birth), sex (women vs men), ethnicity (white vs other ethnic groups), and total non-pension household wealth categorized into quintiles, which represented the combined value of financial assets, physical possessions, and housing owned by a household (consisting of either a single respondent or a responding couple with any dependents), minus any debts.[101] Age completed full-time education was categorized as none, age 14 or under, age 15, age 16, age 17, age 18, and age 19 or over.

Health behaviors included self-reported smoking status, categorized as never, former, or current smoker. Alcohol consumption was self-reported, and categorized as five to seven days a week, one to four days a week, once or twice a month, once or twice a year or once every couple of months, or not at all in the last 12 months or not applicable. Physical activity was reported by the participants, categorized into more than weekly moderate and vigorous exercise, weekly moderate or vigorous exercise, monthly moderate and/or vigorous exercise, or less than monthly moderate and vigorous exercise. We also included several indicators of mental and physical health status: physician-diagnosed cardiovascular disease (heart attack, angina, or heart failure). Body mass index (BMI) was calculated using participant’s height and weight measured during the nurse visit. Depressive symptoms were evaluated using the eight-item Center for Epidemiologic Studies Depression (CESD) scale,[102] in which the participants were asked to indicate, via a binary yes or no response, whether they had experienced symptoms such as restless sleep and sadness during the past week. Scores ranged from 0 to 8, with higher scores indicating more pronounced depressive symptoms.

The APOE genotype was derived from the analysis of two specific SNPs, namely rs7412 and rs429358. To determine these genotypes, two TaqMan assays from Assay-On-Demand, a product of Applied Biosystems and Gene service Ltd in Cambridge, UK, were employed. These assays were conducted on a 7900HT analyzer, manufactured by Applied Biosystems, and the genotypes were determined using the Sequence Detection Software (version 2.0), also from Applied Biosystems. The quality control procedures of genome-wide genotyping has been described elsewhere.[103] The APOE genotype was categorized into a binary variable (APOE ε4 carrier and non-carrier).

### Statistical analysis

#### Missing data handling and imputation

All proteins had <6% missingness. Missing data in the proteomics data and covariates were handled using a random forest imputation method, via the ‘mice’ package in R,[104] which enables the model to account for non-linear relationships and reduce potential biases caused by missing data. Missing values for the cognitive measures were not imputed, as linear mixed-effects models (LMM) can manage missing outcome data through their robust handling of incomplete longitudinal records.[105] A single imputed dataset with 10 iterations was created.

#### Main analysis: Mixed-effect linear regression for proteins associated with cognitive trajectories

To examine the associations of protein levels on the rate of cognitive decline, we used LMM constructed with the ‘lmer’ function from the ‘lme4’ and ‘lmerTest’ packages in R.[106, 107] Models were estimated separately for each protein. These models assessed how the protein levels measured at wave 4 were associated with cognitive trajectories over the following 15 years. The models included random intercepts and slopes for each individual to account for both within-subject variability in cognition at baseline and the longitudinal structure of the data. Models allowed for an interaction between protein level and time since protein measurement and adjusted for baseline age, sex, ethnicity, education, wealth quintile for minimal adjustments, and further adjusted for self-reported cardiovascular disease, diabetes, stroke, BMI, systolic blood pressure, depressive symptoms, smoking status, physical activity, and alcohol consumption for full adjustments. The two-way interaction term (protein × time) was used to report the effect on the rate of change in cognitive function over time, with false discovery rate (FDR) correction applied.

#### Subgroup analysis: Mixed-effect linear regression by sex, APOE ε4 carriage, and age at baseline

Cognitive trajectories were modeled as a function of age using fully adjusted LMMs by subgroups (sex, age at baseline, APOE ε4 carriage). To evaluate subgroup differences in age-related cognitive change, we tested interaction terms between age and the subgroup of interest. To further illustrate the estimated age-related cognitive change in each subgroup, predicted marginal trajectories stratified by subgroup were estimated using the ‘ggpredict’ function in the ‘ggeffects’ package[108] in R, with 95% confidence intervals plotted to visualize uncertainty in the age-by-subgroup interaction effects.

Subgroup stratified analyses for protein associated with cognitive trajectory were further conducted, including by sex (women vs men), APOE ε4 carriage (no copy of APOE ε4 allele vs at least one copy of APOE ε4 allele), and age at baseline (≤65 vs >65 years), using the same model specification as the LMMs in the main analysis.

#### Enrichment network analysis

Enrichment network analysis was performed by querying the following databases using EnrichR[109], using all identified proteins from the main and subgroup, separated according to cognitive domains: The Gene Ontology (GO) database categorizes genes into biological processes, molecular functions, and cellular components, facilitating the understanding of protein function and localization within biological systems;[110] The Kyoto Encyclopedia of Genes and Genomes (KEGG) database, which offers a collection of pathways and functional annotations, allowing for the identification of biological pathways enriched;[111] The REACTOME database, which contains curated pathways and reactions involved in various biological processes, enabling the exploration of molecular events and signaling pathways.[112]

The gene set libraries from EnrichR-KG[109, 113, 114] were transformed into a network of nodes and links, where nodes represent either gene set terms or individual genes. This process generated a visual representation of the relationships between enriched terms and genes across multiple annotated gene set libraries, leveraging existing biological knowledge. By constructing an integrated gene network, enrichment analysis was conducted to identify biological pathways and functional annotations related to the input gene list in EnrichR-KG (https://maayanlab.cloud/enrichr-kg). The top ten terms with P<0.05 from each library were included. Data visualization for the network analysis was then performed using the ‘NetworkX’ library[115] in Python.

To further enhance the enrichment network analysis, protein expression patterns and functional annotations were cross-referenced using the Human Protein Atlas,[33] thereby providing additional biological context. Additional enrichment analyses were conducted to characterize the biological relevance of proteins passing the nominal significance threshold (P_FDR_ < 0.05). Gene identifiers were standardized to HGNC-compliant gene symbols prior to downstream analysis. Over-representation analysis was conducted across multiple pathway databases, including GO, KEGG pathways, and Reactome pathways using g:Profiler (gost). Statistical significance was assessed with Benjamini–Hochberg FDR correction, and pathways with adjusted P<0.05 were considered significant. Analyses were restricted to protein-coding genes with valid annotation in the respective databases. Results were visualized as bubble plots showing the top 15 enriched terms, with point size indicating the number of overlapping genes and color intensity indicating statistical significance (−log10 adjusted P-value).

Gene variants, eQTLs and associated biological/clinical annotations for proteins of interest were explored with the following tools and data,[116–118] and the NHGRI-EBI GWAS Catalog: https://www.ebi.ac.uk/gwas/home.[119]

#### Open target for drug identification

To explore potential therapeutic interventions, we queried the Open Targets platform[120] for clinically trialed drugs targeting proteins identified in our main and subgroup analyses. For proteins associated with accelerated cognitive decline, we generated a Sankey diagram to map the relationships between these proteins, their corresponding drugs, and the drugs’ mechanisms of action (e.g., agonists, inhibitors, or binding agents). This visualization was created using the ‘networkD3’ R package[121], providing an intuitive representation of the therapeutic landscape and biological pathways implicated in cognitive decline.

All analyses were performed using RStudio (version 4.5.0) and Jupyter Notebook with Python (version 3.11.7).

## Data Availability

The ELSA data is available on the UK Data Service.

## Acknowledgement

We would like to thank the participants in ELSA for their contribution to the research.

The English Longitudinal Study of Ageing is funded by the National Institute on Aging (grant number R01AG17644) and the National Institute for Health and Care Research (198/1074-02). The National Institute of Aging (NIA) (grant Number [R01AG17644]) funded the proteomics data curation in ELSA. J.G. is supported by the NIA (grant Number [R01AG17644]) and the Wellcome Leap CARE grant.

The funders had no role in study design; in the collection, analysis and interpretation of data; in the writing of the report; and in the decision to submit the article for publication.

## Data availability

The ELSA data is available on the UK Data Service.

## Code availability

The codes used for all analyses are available on GitHub repository: https://github.com/jgong94/ELSA_proteomics_cognition.

## References

1. Livingston, G., et al., Dementia prevention, intervention, and care: 2024 report of the Lancet standing Commission. The Lancet, 2024. 404(10452): p. 572–628.

2. Nichols, E., et al., Estimation of the global prevalence of dementia in 2019 and forecasted prevalence in 2050: an analysis for the Global Burden of Disease Study 2019. The Lancet Public Health, 2022. 7(2): p. e105–e125.

3. Wang, X., et al., *Blood*□*based biomarkers for Alzheimer’s disease and cognitive function from mid*□*to late life*. Alzheimer’s & Dementia, 2024. 20(3): p. 1807–1814.

4. Organization, W.H., Risk reduction of cognitive decline and dementia: WHO guidelines. 2019: World Health Organization.

5. Sun, B.B., K. Suhre, and B.W. Gibson, Promises and challenges of populational proteomics in health and disease. Molecular & Cellular Proteomics, 2024: p. 100786.

6. Gong, J., et al., Unraveling the role of plasma proteins in dementia: insights from two cohort studies in the UK, with causal evidence from Mendelian randomization (in press). Brain Communications, 2025: p. 2024.06.04.24308415.

7. Lindbohm, J.V., et al., *Plasma proteins, cognitive decline, and 20*□*year risk of dementia in the Whitehall II and Atherosclerosis Risk in Communities studies*. Alzheimer’s & dementia, 2022. 18(4): p. 612–624.

8. Walker, K.A., et al., Proteomics analysis of plasma from middle-aged adults identifies protein markers of dementia risk in later life. Science translational medicine, 2023. 15(705): p. eadf5681.

9. Walker, K.A., et al., Large-scale plasma proteomic analysis identifies proteins and pathways associated with dementia risk. Nature Aging, 2021. 1(5): p. 473–489.

10. Guo, Y., et al., Plasma proteomic profiles predict future dementia in healthy adults. Nature Aging, 2024. 4(2): p. 247–260.

11. Trares, K., et al., Association of the inflammation-related proteome with dementia development at older age: results from a large, prospective, population-based cohort study. Alzheimer’s Research & Therapy, 2022. 14(1): p. 128.

12. Deng, Y.-T., et al., Atlas of the plasma proteome in health and disease in 53,026 adults. Cell, 2024.

13. Harris, S.E., et al., Neurology-related protein biomarkers are associated with cognitive ability and brain volume in older age. Nature Communications, 2020. 11(1): p. 800.

14. Llaurador-Coll, M., et al., Plasma levels of neurology-related proteins are associated with cognitive performance in an older population with overweight/obesity and metabolic syndrome. GeroScience, 2023. 45(4): p. 2457–2470.

15. Chen, J., et al., Peripheral inflammatory biomarkers are associated with cognitive function and dementia: Framingham Heart Study Offspring cohort. Aging cell, 2023. 22(10): p. e13955.

16. Zheng, Y., et al., Exploring the relationship between lipid metabolism and cognition in individuals living with stable-phase Schizophrenia: a small cross-sectional study using Olink proteomics analysis. BMC psychiatry, 2024. 24(1): p. 593.

17. Tanaka, T., et al., Plasma proteomic signatures predict dementia and cognitive impairment. Alzheimers. Dement. 6, e12018. 2020.

18. Kresge, H.A., et al., *Discovery*□*based proteomics identifies plasma proteins that predict longitudinal cognitive decline in older adults over a 7*□*year follow*□*up period*. Alzheimer’s & Dementia, 2023. 19: p. e079341.

19. Duggan, M.R., et al., Proteomics identifies potential immunological drivers of postinfection brain atrophy and cognitive decline. Nature Aging, 2024. 4(9): p. 1263–1278.

20. Kivisäkk, P., et al., Plasma biomarkers for prognosis of cognitive decline in patients with mild cognitive impairment. Brain Communications, 2022. 4(4): p. fcac155.

21. Duggan, M.R., et al., Plasma proteins related to inflammatory diet predict future cognitive impairment. Molecular psychiatry, 2023. 28(4): p. 1599–1609.

22. Rabl, M., et al., *Neuropsychiatric symptoms in cognitive decline and Alzheimer’s disease: biomarker discovery using plasma proteomics.* Journal of Neurology, Neurosurgery & Psychiatry, 2025. 96(4): p. 370–382.

23. Sim, M.A., et al., Plasma proteomics for cognitive decline and dementia—A Southeast Asian cohort study. Alzheimer’s & dementia, 2025. 21(2): p. e14577.

24. Adegboye, H.A., et al., *LC*□*MS/MS proteomics identifies plasma proteins related to cognition over 9*□*year follow*□*up*. Alzheimer’s & Dementia, 2025. 21(6): p. e70276.

25. Duggan, M.R., et al., Proteomics identifies potential immunological drivers of postinfection brain atrophy and cognitive decline. Nature Aging, 2024: p. 1–16.

26. Tanaka, T., et al., Plasma proteomic signatures predict dementia and cognitive impairment. Alzheimer’s & Dementia: Translational Research & Clinical Interventions, 2020. 6(1): p. e12018.

27. Nagy, D., et al., Developmental synaptic regulator, TWEAK/Fn14 signaling, is a determinant of synaptic function in models of stroke and neurodegeneration. Proceedings of the National Academy of Sciences, 2021. 118(6): p. e2001679118.

28. Jéru, I., et al., Brief report: involvement of TNFRSF11A molecular defects in autoinflammatory disorders. Arthritis & Rheumatology, 2014. 66(9): p. 2621–2627.

29. Kaczynski, T.J., et al., Dysregulation of a lncRNA within the TNFRSF10A locus activates cell death pathways. Cell Death Discovery, 2023. 9(1): p. 242.

30. Fourgeaud, L., et al., TAM receptors regulate multiple features of microglial physiology. Nature, 2016. 532(7598): p. 240–244.

31. Huang, Y., et al., *Microglia use TAM receptors to detect and engulf amyloid* β *plaques*. Nature immunology, 2021. 22(5): p. 586–594.

32. Hu, Z., et al., Novel inflammatory markers in intracerebral hemorrhage: Results from Olink proteomics analysis. The FASEB Journal, 2025. 39(2): p. e70341.

33. Uhlen, M., et al., Towards a knowledge-based human protein atlas. Nature biotechnology, 2010. 28(12): p. 1248–1250.

34. Angiari, S., et al., TIM-1 glycoprotein binds the adhesion receptor P-selectin and mediates T cell trafficking during inflammation and autoimmunity. Immunity, 2014. 40(4): p. 542–553.

35. Yirmiya, R. and I. Goshen, Immune modulation of learning, memory, neural plasticity and neurogenesis. Brain, behavior, and immunity, 2011. 25(2): p. 181–213.

36. Heneka, M.T., et al., Neuroinflammation in Alzheimer’s disease. The Lancet Neurology, 2015. 14(4): p. 388–405.

37. Ljungqvist, J., et al., Serum neurofilament light protein as a marker for diffuse axonal injury: results from a case series study. Journal of neurotrauma, 2017. 34(5): p. 1124–1127.

38. Graham, N.S., et al., Axonal marker neurofilament light predicts long-term outcomes and progressive neurodegeneration after traumatic brain injury. Science translational medicine, 2021. 13(613): p. eabg9922.

39. Zetterberg, H., Neurofilament light: a dynamic cross-disease fluid biomarker for neurodegeneration. Neuron, 2016. 91(1): p. 1–3.

40. Gaetani, L., et al., *Neurofilament light chain as a biomarker in neurological disorders.* Journal of Neurology, Neurosurgery & Psychiatry, 2019. 90(8): p. 870–881.

41. Olsson, B., et al., Association of cerebrospinal fluid neurofilament light protein levels with cognition in patients with dementia, motor neuron disease, and movement disorders. JAMA neurology, 2019. 76(3): p. 318–325.

42. Ashton, N.J., et al., A multicentre validation study of the diagnostic value of plasma neurofilament light. Nature communications, 2021. 12(1): p. 3400.

43. Disanto, G., et al., Serum neurofilament light: a biomarker of neuronal damage in multiple sclerosis. Annals of neurology, 2017. 81(6): p. 857–870.

44. Salvadores, N., C. Gerónimo-Olvera, and F.A. Court, *Axonal degeneration in AD: the contribution of A*β *and Tau*. Frontiers in Aging Neuroscience, 2020. 12: p. 581767.

45. Held, F., et al., Proteomics reveals age as major modifier of inflammatory CSF signatures in multiple sclerosis. Neurology: Neuroimmunology & Neuroinflammation, 2024. 12(1): p. e200322.

46. Lin, J., J. Zhou, and Y. Xu, Potential drug targets for multiple sclerosis identified through Mendelian randomization analysis. Brain, 2023. 146(8): p. 3364–3372.

47. Philippi, S.M., et al., APOE genotype and brain amyloid are associated with changes in the plasma proteome in elderly subjects without dementia. Annals of Clinical and Translational Neurology, 2025. 12(2): p. 366–382.

48. Li, J., et al., Cryo-EM analyses reveal the common mechanism and diversification in the activation of RET by different ligands. Elife, 2019. 8: p. e47650.

49. Shao, Q., et al., Uncoupling of UNC5C with polymerized TUBB3 in microtubules mediates netrin-1 repulsion. Journal of Neuroscience, 2017. 37(23): p. 5620–5633.

50. Montagne, A., et al., Blood-brain barrier breakdown in the aging human hippocampus. Neuron, 2015. 85(2): p. 296–302.

51. Hartz, A.M., et al., *Amyloid-*β *contributes to blood–brain barrier leakage in transgenic human amyloid precursor protein mice and in humans with cerebral amyloid angiopathy*. Stroke, 2012.

52. Yang, H.-S., et al., Plasma VEGFA and PGF impact longitudinal tau and cognition in preclinical Alzheimer’s disease. Brain, 2024. 147(6): p. 2158–2168.

53. Gunstad, J., et al., Relation of brain natriuretic peptide levels to cognitive dysfunction in adults> 55 years of age with cardiovascular disease. The American journal of cardiology, 2006. 98(4): p. 538–540.

54. Kerola, T., et al., B-type natriuretic peptide as a predictor of declining cognitive function and dementia—a cohort study of an elderly general population with a 5-year follow-up. Annals of medicine, 2010. 42(3): p. 207–215.

55. van Vliet, P., et al., NT-proBNP, blood pressure, and cognitive decline in the oldest old: The Leiden 85-plus Study. Neurology, 2014. 83(13): p. 1192–1199.

56. Daniels, L.B., et al., Elevated natriuretic peptide levels and cognitive function in community-dwelling older adults. The American Journal of Medicine, 2011. 124(7): p. 670. e1–670. e8.

57. Lu, X.-D., Y.-R. Liu, and Z.-Y. Zhang, Matrilin-3 alleviates extracellular matrix degradation of nucleus pulposus cells via induction of IL-1 receptor antagonist. European Review for Medical & Pharmacological Sciences, 2020. 24(10).

58. Zhang, W., et al., Decorin is a pivotal effector in the extracellular matrix and tumour microenvironment. Oncotarget, 2018. 9(4): p. 5480.

59. Hong, R., et al., PRELP has prognostic value and regulates cell proliferation and migration in hepatocellular carcinoma. Journal of Cancer, 2020. 11(21): p. 6376.

60. Lucarelli, G., et al., Spondin-2, a secreted extracellular matrix protein, is a novel diagnostic biomarker for prostate cancer. The Journal of urology, 2013. 190(6): p. 2271–2277.

61. Maier, S., M. Paulsson, and U. Hartmann, The widely expressed extracellular matrix protein SMOC-2 promotes keratinocyte attachment and migration. Experimental cell research, 2008. 314(13): p. 2477–2487.

62. Arrate, M.P., et al., Cloning of human junctional adhesion molecule 3 (JAM3) and its identification as the JAM2 counter-receptor. Journal of Biological Chemistry, 2001. 276(49): p. 45826–45832.

63. Rosenberg, G.A., Extracellular matrix inflammation in vascular cognitive impairment and dementia. Clinical science, 2017. 131(6): p. 425–437.

64. Tisato, V., et al., Clinical perspectives of TRAIL: insights into central nervous system disorders. Cellular and Molecular Life Sciences, 2016. 73: p. 2017–2027.

65. Di Benedetto, G., et al., *TRAIL-R deficient mice are protected from neurotoxic effects of amyloid-*β. International Journal of Molecular Sciences, 2022. 23(19): p. 11625.

66. Palomer, E., J. Buechler, and P.C. Salinas, Wnt signaling deregulation in the aging and Alzheimer’s brain. Frontiers in cellular neuroscience, 2019. 13: p. 227.

67. Inestrosa, N.C. and L. Varela-Nallar, Wnt signaling in the nervous system and in Alzheimer’s disease. Journal of molecular cell biology, 2014. 6(1): p. 64–74.

68. Liu, W.Y., et al., *Tight junction in blood*□*brain barrier: an overview of structure, regulation, and regulator substances*. CNS neuroscience & therapeutics, 2012. 18(8): p. 609–615.

69. Bowman, G.L., et al., Blood-brain barrier breakdown, neuroinflammation, and cognitive decline in older adults. Alzheimer’s & Dementia, 2018. 14(12): p. 1640–1650.

70. Owens, C.D., et al., Microvascular dysfunction and neurovascular uncoupling are exacerbated in peripheral artery disease, increasing the risk of cognitive decline in older adults. American Journal of Physiology-Heart and Circulatory Physiology, 2022. 322(6): p. H924–H935.

71. Ruan, Z., et al., Altered neurovascular coupling in patients with vascular cognitive impairment: a combined ASL-fMRI analysis. Frontiers in Aging Neuroscience, 2023. 15: p. 1224525.

72. Stuss, D.T. and M.P. Alexander, Executive functions and the frontal lobes: a conceptual view. Psychological research, 2000. 63(3): p. 289–298.

73. Shomstein, S., Cognitive functions of the posterior parietal cortex: top-down and bottom-up attentional control. Frontiers in integrative neuroscience, 2012. 6: p. 38.

74. Tragantzopoulou, P. and V. Giannouli, Spatial orientation assessment in the elderly: a comprehensive review of current tests. Brain Sciences, 2024. 14(9): p. 898.

75. Qian, X.-H., et al., Blood Cathepsins on the Risk of Alzheimer’s Disease and Related Pathological Biomarkers: Results from Observational Cohort and Mendelian Randomization Study. The Journal of Prevention of Alzheimer’s Disease, 2024. 11(6): p. 1834–1842.

76. Yamaguchi, N., et al., Spinesin/TMPRSS5, a novel transmembrane serine protease, cloned from human spinal cord. Journal of Biological Chemistry, 2002. 277(9): p. 6806–6812.

77. Liu, K., et al., Transferrin receptor controls AMPA receptor trafficking efficiency and synaptic plasticity. Scientific reports, 2016. 6(1): p. 21019.

78. Hémar, A., et al., Dendroaxonal transcytosis of transferrin in cultured hippocampal and sympathetic neurons. Journal of Neuroscience, 1997. 17(23): p. 9026–9034.

79. Morris, C., et al., Transferrin receptors in the normal human hippocampus and in Alzheimer’s disease. Neuropathology and applied neurobiology, 1994. 20(5): p. 473–477.

80. Zhang, Y., et al., Hippocampal iron accumulation impairs synapses and memory via suppressing furin expression and downregulating BDNF maturation. Molecular Neurobiology, 2022. 59(9): p. 5574–5590.

81. Carlson, E.S., et al., Iron is essential for neuron development and memory function in mouse hippocampus. The Journal of nutrition, 2009. 139(4): p. 672–679.

82. Ferreri, N.R., Estrogen-TNF interactions and vascular inflammation. American Journal of Physiology-Heart and Circulatory Physiology, 2007. 292(6): p. H2566–H2569.

83. Xing, D., et al., *Estrogen modulates TNF-*α*-induced inflammatory responses in rat aortic smooth muscle cells through estrogen receptor-*β *activation*. American Journal of Physiology-Heart and Circulatory Physiology, 2007. 292(6): p. H2607–H2612.

84. Oh, H.S.-H., et al., Plasma proteomics links brain and immune system aging with healthspan and longevity. Nature Medicine, 2025: p. 1–9.

85. Klein, S.L. and K.L. Flanagan, Sex differences in immune responses. Nature Reviews Immunology, 2016. 16(10): p. 626–638.

86. Bianchi, I., et al., The X chromosome and immune associated genes. Journal of autoimmunity, 2012. 38(2-3): p. J187–J192.

87. Underwood, B.R., et al., *Data*□*driven discovery of associations between prescribed drugs and dementia risk: A systematic review*. Alzheimer’s & Dementia: Translational Research & Clinical Interventions, 2025. 11(1): p. e70037.

88. Yue, Q., et al., Endothelial Dysfunctions in Blood–Brain Barrier Breakdown in Alzheimer’s Disease: From Mechanisms to Potential Therapies. CNS Neuroscience & Therapeutics, 2024. 30(11): p. e70079.

89. Kim, I.-d., J.W. Cave, and S. Cho, Aflibercept, a VEGF (vascular endothelial growth factor)-trap, reduces vascular permeability and stroke-induced brain swelling in obese mice. Stroke, 2021. 52(8): p. 2637–2648.

90. Campbell, K.S., A.D. Cohen, and T. Pazina, Mechanisms of NK cell activation and clinical activity of the therapeutic SLAMF7 antibody, elotuzumab in multiple myeloma. Frontiers in immunology, 2018. 9: p. 2551.

91. Merchant, M.S., et al., Phase I trial and pharmacokinetic study of lexatumumab in pediatric patients with solid tumors. Journal of clinical oncology, 2012. 30(33): p. 4141–4147.

92. Zhang, L., et al., Lexatumumab (TRAIL-receptor 2 mAb) induces expression of DR5 and promotes apoptosis in primary and metastatic renal cell carcinoma in a mouse orthotopic model. Cancer letters, 2007. 251(1): p. 146–157.

93. MacAulay, R.K., et al., Understanding heterogeneity in older adults: Latent growth curve modeling of cognitive functioning. Journal of clinical and experimental neuropsychology, 2018. 40(3): p. 292–302.

94. Heitz, R.P., et al., Working memory, executive function, and general fluid intelligence are not the same. Behavioral and Brain Sciences, 2006. 29(2): p. 135–136.

95. Hamer, M., G.M. Terrera, and P. Demakakos, Physical activity and trajectories in cognitive function: English Longitudinal Study of Ageing. J Epidemiol Community Health, 2018. 72(6): p. 477–483.

96. Uhlén, M., et al., Tissue-based map of the human proteome. Science, 2015. 347(6220): p. 1260419.

97. Zheng, F., et al., HbA1c, diabetes and cognitive decline: the English Longitudinal Study of Ageing. Diabetologia, 2018. 61(4): p. 839–848.

98. Zaninotto, P., et al., Cognitive function trajectories and their determinants in older people: 8 years of follow-up in the English Longitudinal Study of Ageing. J Epidemiol Community Health, 2018. 72(8): p. 685–694.

99. Steptoe, A., et al., Cohort profile: the English longitudinal study of ageing. International journal of epidemiology, 2013. 42(6): p. 1640–1648.

100. Chertkow, H., et al., Mild cognitive impairment and cognitive impairment, no dementia: Part A, concept and diagnosis. Alzheimer’s & Dementia, 2007. 3(4): p. 266–282.

101. Zaninotto, P. and C. Lassale, Socioeconomic trajectories of body mass index and waist circumference: results from the English Longitudinal Study of Ageing. BMJ open, 2019. 9(4): p. e025309.

102. Radloff, L.S., The use of the Center for Epidemiologic Studies Depression Scale in adolescents and young adults. Journal of youth and adolescence, 1991. 20(2): p. 149–166.

103. Ajnakina, O., D. Cadar, and A. Steptoe, Interplay between socioeconomic markers and polygenic predisposition on timing of dementia diagnosis. Journal of the American Geriatrics Society, 2020. 68(7): p. 1529–1536.

104. Van Buuren, S. and K. Groothuis-Oudshoorn, mice: Multivariate imputation by chained equations in R. Journal of statistical software, 2011. 45: p. 1–67.

105. Chhabra, G., V. Vashisht, and J. Ranjan, A comparison of multiple imputation methods for data with missing values. Indian Journal of Science and Technology, 2017. 10(19): p. 1–7.

106. Bates, D., et al., Package ‘lme4’. convergence, 2015. 12(1): p. 2.

107. Kuznetsova, A., P.B. Brockhoff, and R.H. Christensen, lmerTest package: tests in linear mixed effects models. Journal of statistical software, 2017. 82: p. 1–26.

108. Lüdecke, D., *ggeffects: Tidy data frames of marginal effects from regression models*. Journal of open source software, 2018. 3(26): p. 772.

109. Xie, Z., et al., Gene set knowledge discovery with Enrichr. Current protocols, 2021. 1(3): p. e90.

110. Consortium, G.O., The Gene Ontology (GO) database and informatics resource. Nucleic acids research, 2004. 32(suppl_1): p. D258–D261.

111. Kanehisa, M. and S. Goto, KEGG: kyoto encyclopedia of genes and genomes. Nucleic acids research, 2000. 28(1): p. 27–30.

112. Gillespie, M., et al., The reactome pathway knowledgebase 2022. Nucleic acids research, 2022. 50(D1): p. D687–D692.

113. Chen, E.Y., et al., Enrichr: interactive and collaborative HTML5 gene list enrichment analysis tool. BMC bioinformatics, 2013. 14: p. 1–14.

114. Kuleshov, M.V., et al., Enrichr: a comprehensive gene set enrichment analysis web server 2016 update. Nucleic acids research, 2016. 44(W1): p. W90–W97.

115. Hagberg, A., P.J. Swart, and D.A. Schult, Exploring network structure, dynamics, and function using NetworkX. 2008, Los Alamos National Laboratory (LANL), Los Alamos, NM (United States).

116. Graham, A.C., et al., Human longevity and Alzheimer’s disease variants act via microglia and oligodendrocyte gene networks. Brain, 2025. 148(3): p. 969–984.

117. Consortium, G., et al., The Genotype-Tissue Expression (GTEx) pilot analysis: multitissue gene regulation in humans. Science, 2015. 348(6235): p. 648–660.

118. Bryois, J., et al., Cell-type-specific cis-eQTLs in eight human brain cell types identify novel risk genes for psychiatric and neurological disorders. Nature neuroscience, 2022. 25(8): p. 1104–1112.

119. Cerezo, M., et al., The NHGRI-EBI GWAS Catalog: standards for reusability, sustainability and diversity. Nucleic acids research, 2025. 53(D1): p. D998–D1005.

120. Koscielny, G., et al., Open Targets: a platform for therapeutic target identification and validation. Nucleic acids research, 2017. 45(D1): p. D985–D994.

121. Allaire, J., et al., Package ‘networkD3’. D3 JavaScript network graphs from R, 2017.

