## Supplementary Figure for "Proteomics signatures associated with cognitive trajectories: evidence from the English Longitudinal Study of Ageing"

**Supplementary Figure 1. Schematic diagram of participant selection in ELSA proteomics study.**

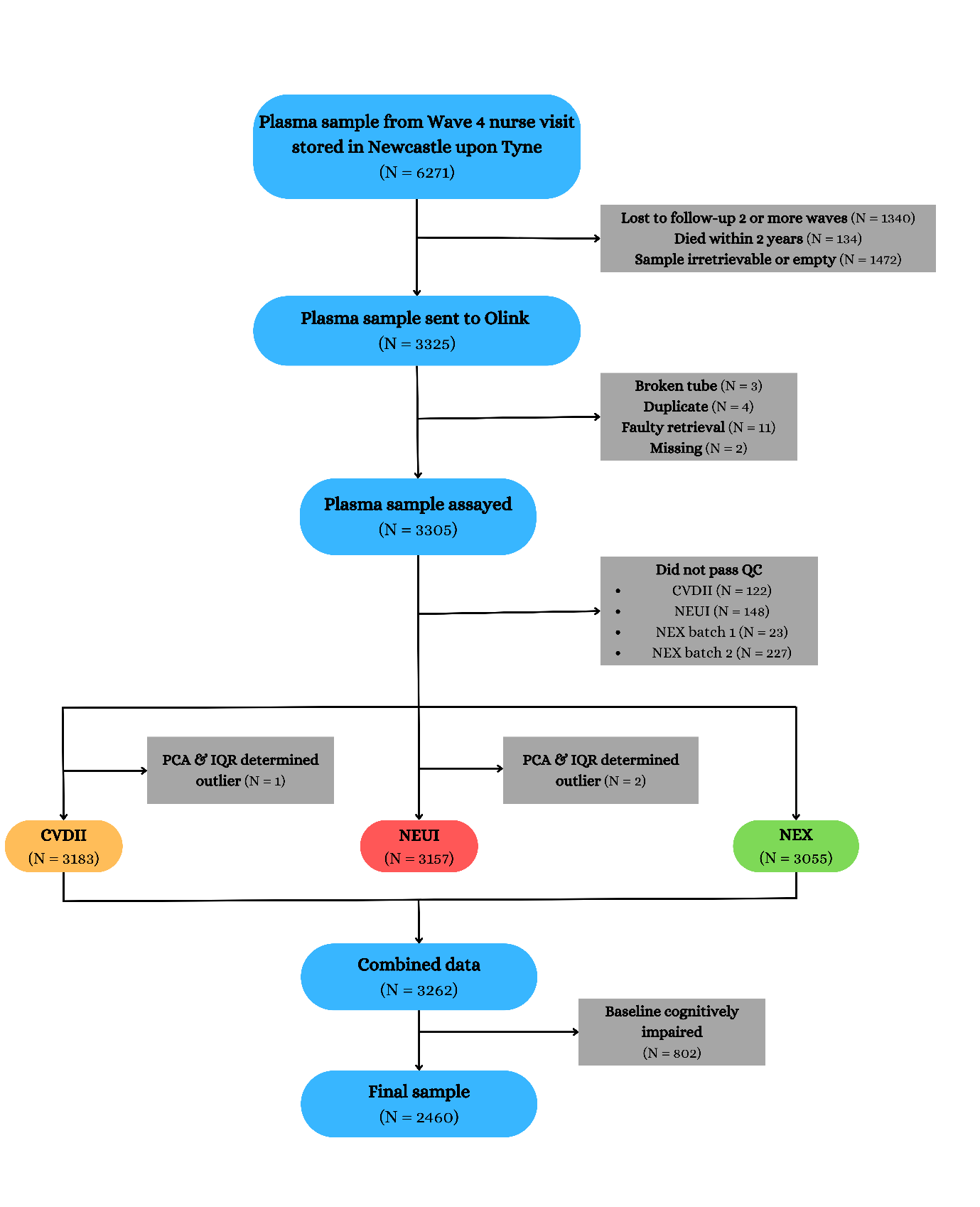

QC, quality control; PCA, principal component analysis; IQR, interquartile range; CVDII, cardiovascular II Olink Target 96 panel; NEUI, neurology I Olink Target 96 panel; NEX, neuro exploratory Olink Target 96 panel.

**Supplementary Figure 2. Volcano plots visualizing protein concentration associated with verbal fluency, episodic memory, orientation in time, from minimally adjusted mixed effect linear regression models.**

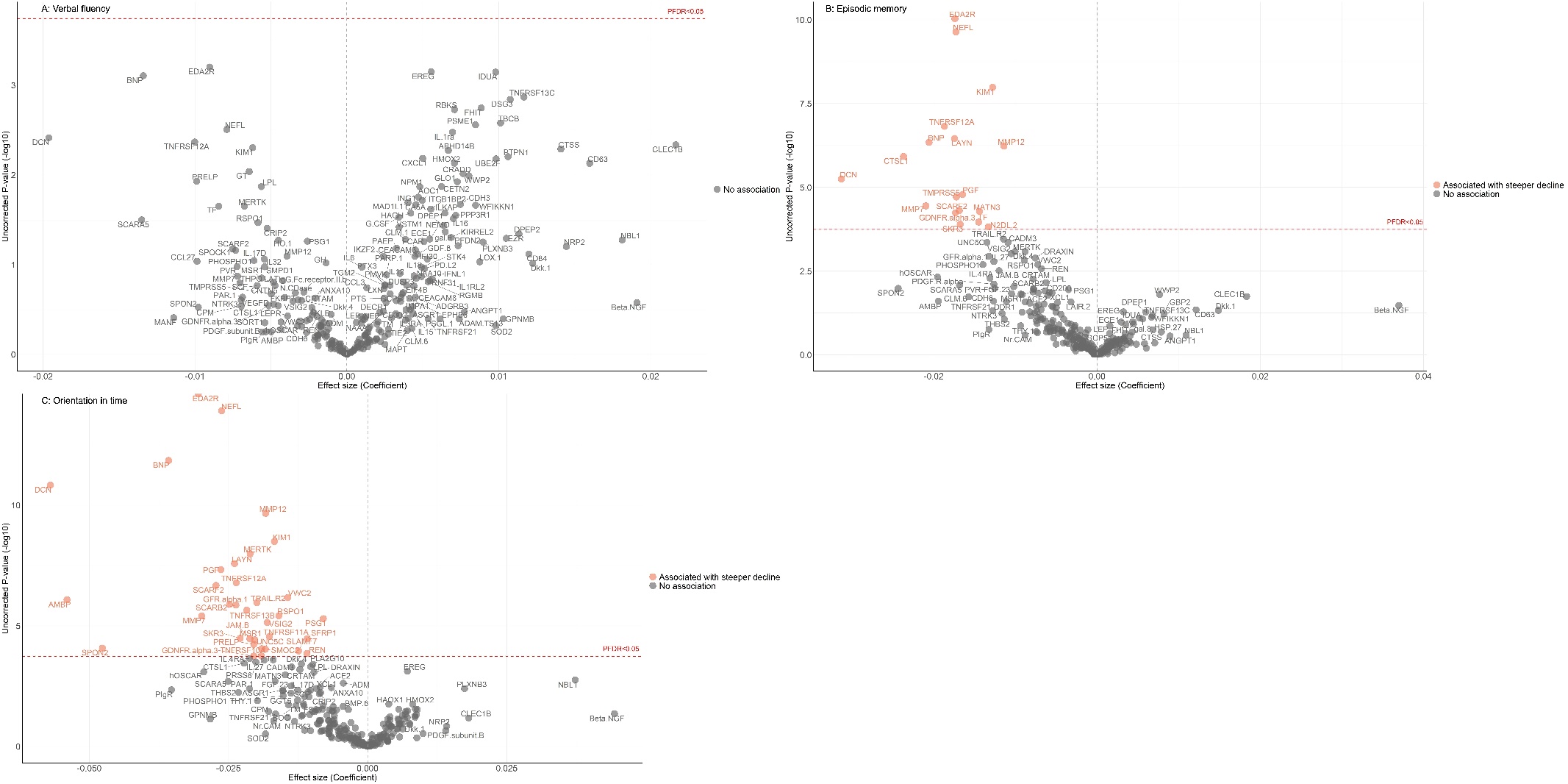

Adjusted for baseline age, sex, ethnicity, education, wealth quintile, allowing for interactions between protein × time since protein measurement.

**Supplementary Figure 3. Predicted cognitive trajectories across age, by sex (women and men).**

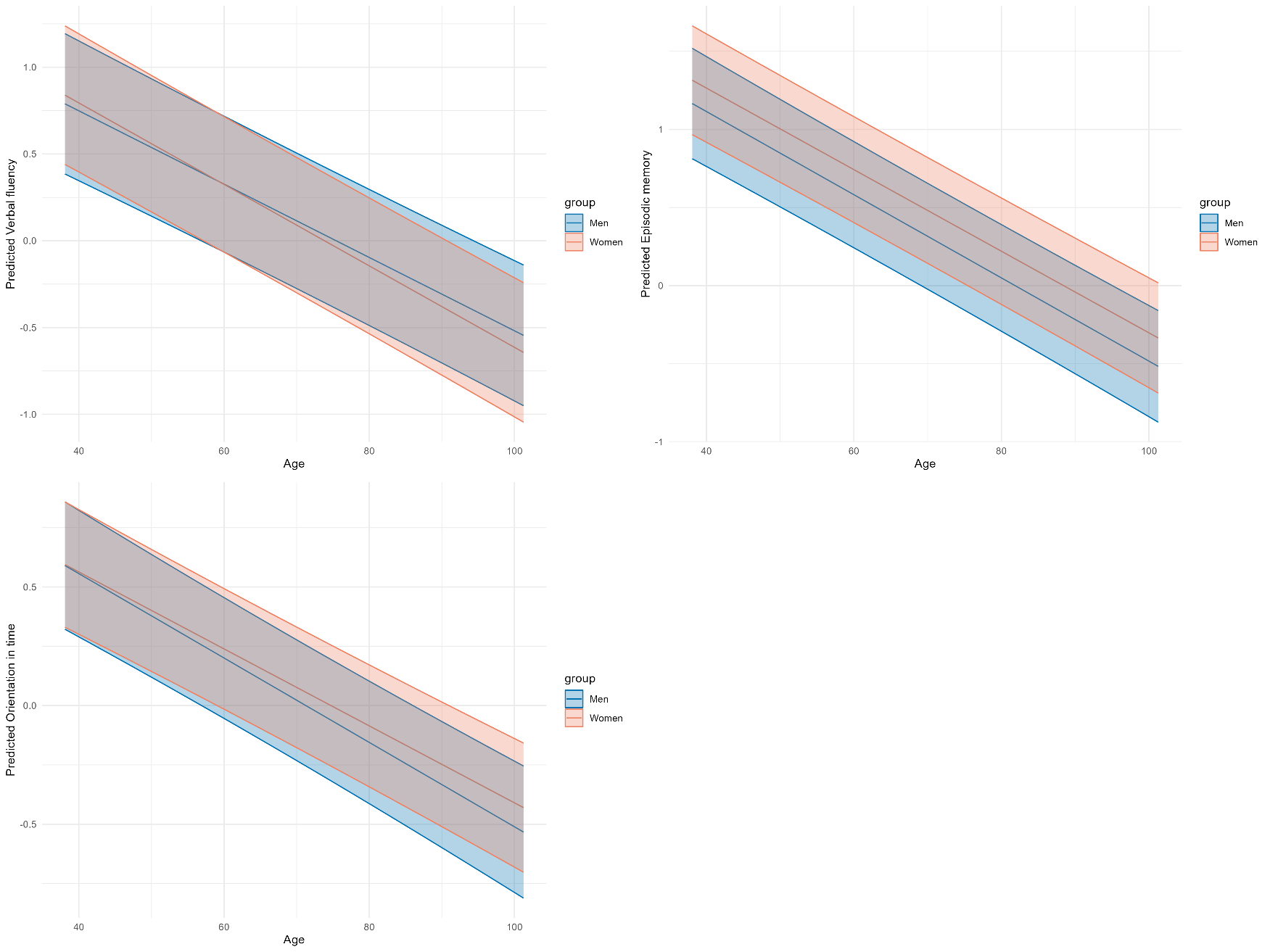

**Supplementary Figure 4. Predicted cognitive trajectories across age, by APOE ɛ4 carriage (carrier and non-carrier).**

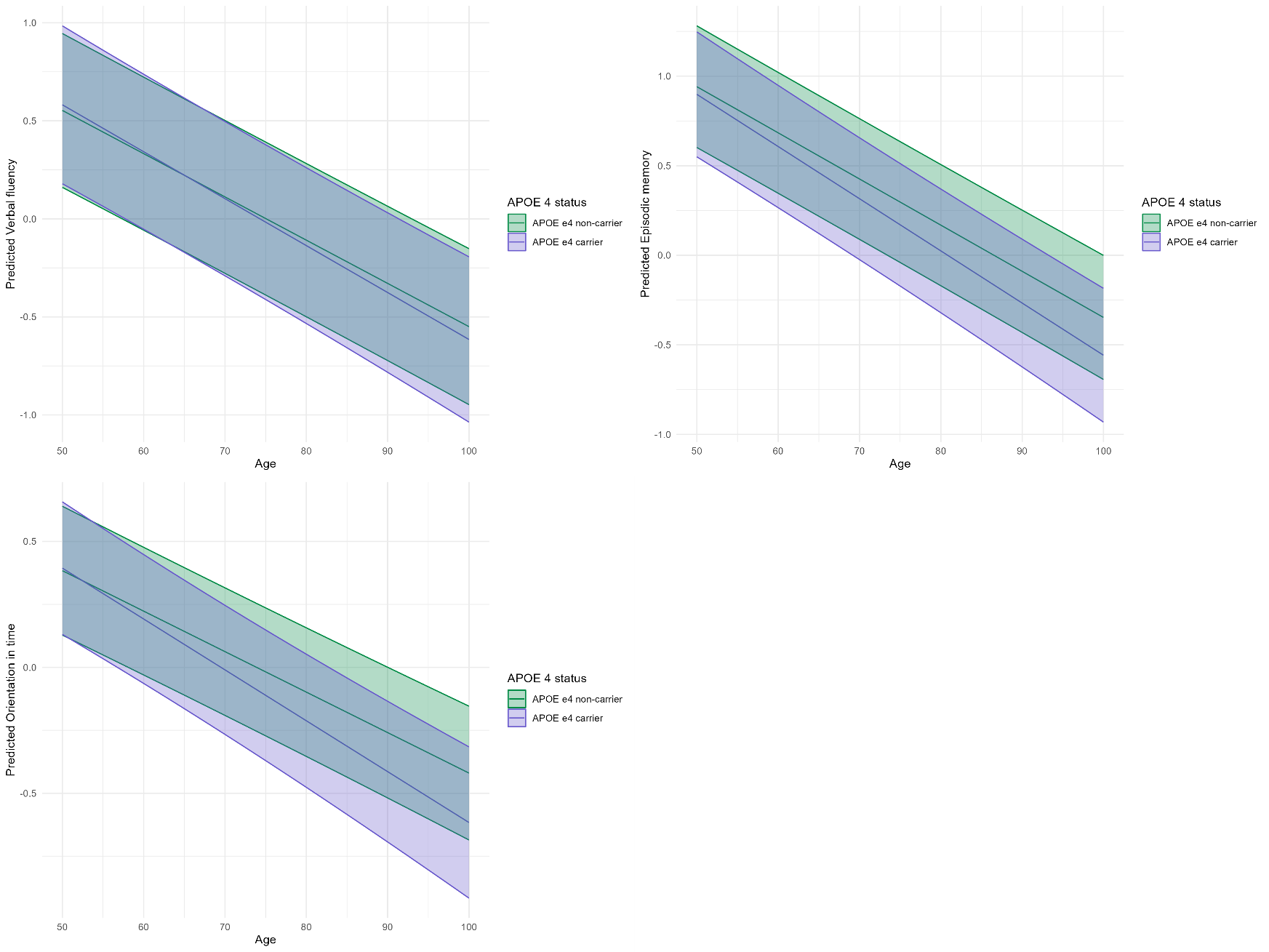

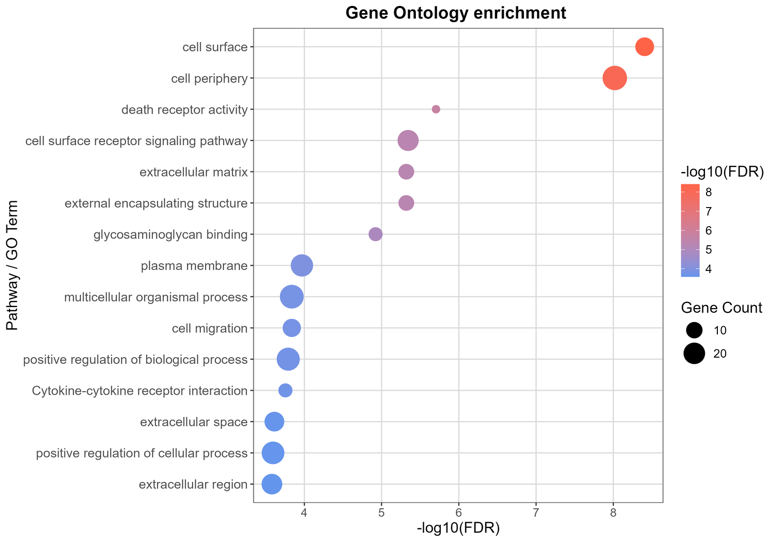
**Supplementary Figure 5. Biological annotations associated with cognitive decline in orientation.**

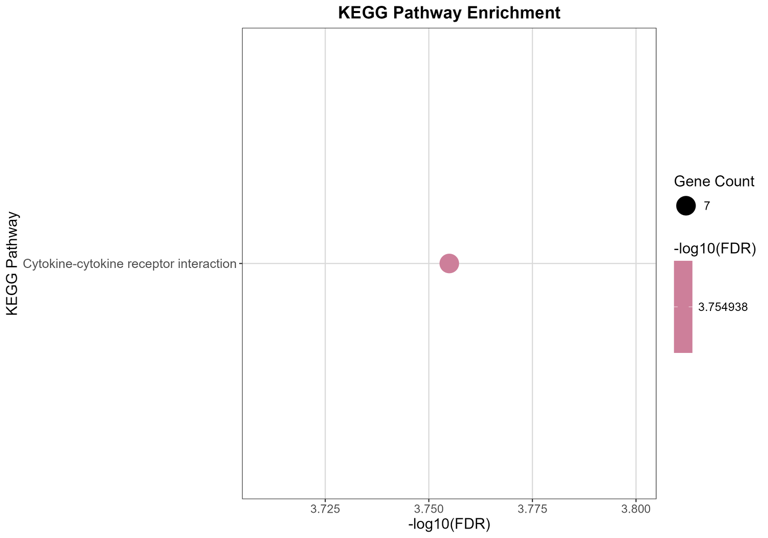

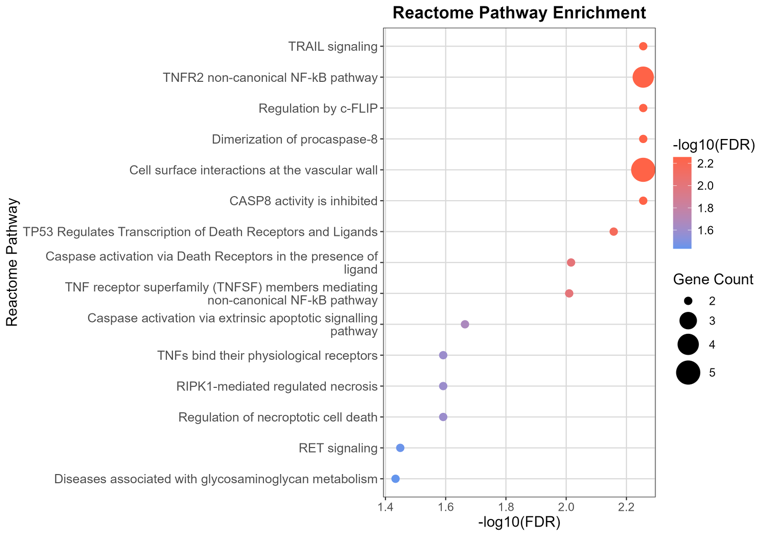

Gene Ontology (GO), KEGG pathway and Reactome pathway enrichment analysis of orientation-associated proteins. Up to the top 15 enriched terms for genes encoding statistically significant proteins (FDR < 0.05) are shown, with dot colour and size representing −log₁₀ FDR and gene count respectively. Enrichment analysis was performed using gost(gprofiler2)[1] with FDR correction against the Homo sapiens reference genome.

**Supplementary Figure 6. Network annotation associated with cognitive decline in orientation.**

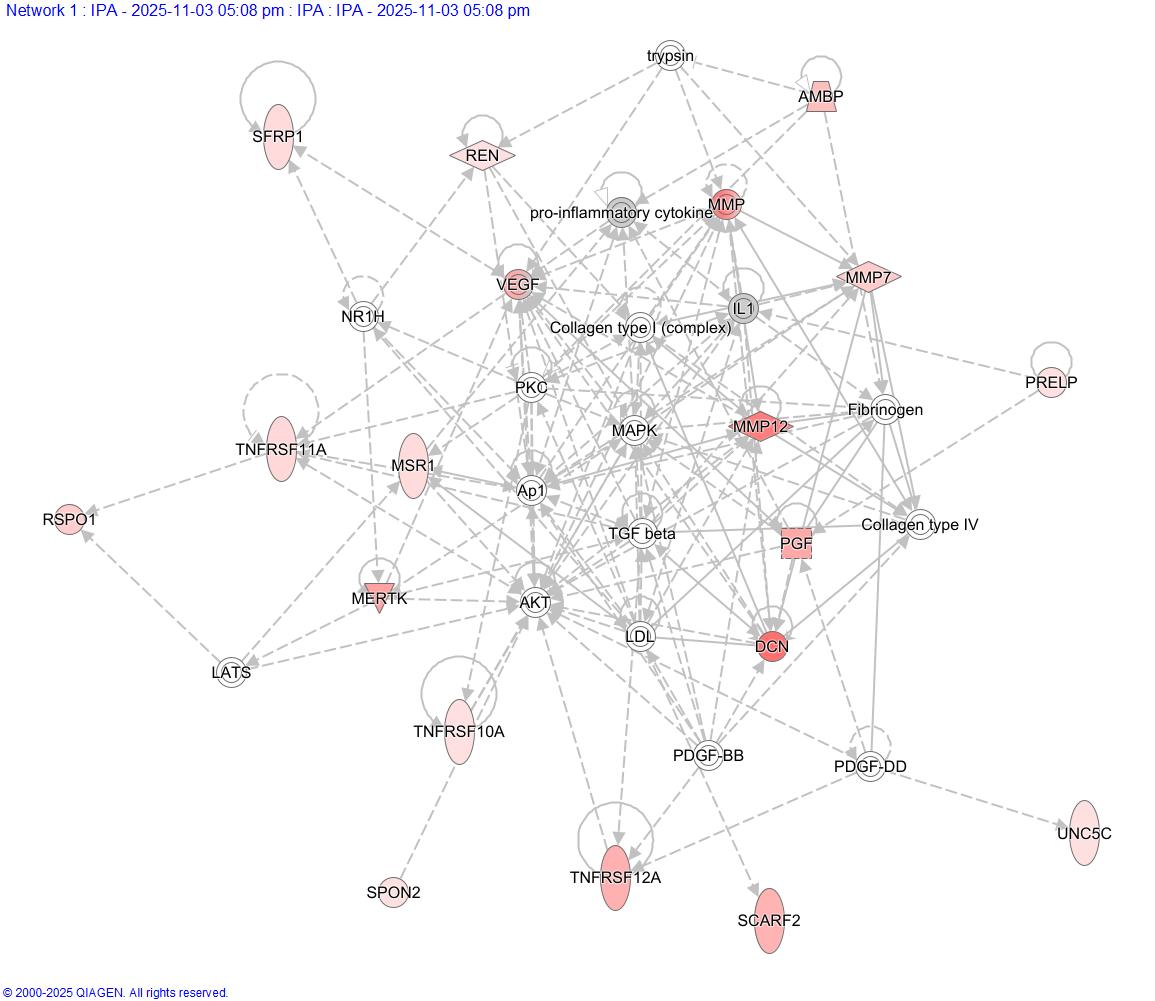

Ingenuity plot showing the proteins associated with cognitive decline in orientation are linked in a network containing TNF signalling and ECM processes focussed on the mitogen-activated protein kinase (MAPK) and transforming growth factor-β (TGF-β) signalling pathways. Solid lines indicate direct interactions (e.g. protein-protein interactions, or phosphorylation), and broken lines indirect interactions, between pairs of genes across all mammalian species for tissues and cell types curated in Ingenuity Pathway Analysis (IPA; QIAGEN). Statistically significant proteins (FDR < 0.05) associated with decline in orientation were used as input. Default settings used.

**Supplementary Figure 7. Top 5 canonical pathways identified by IPA associated with cognitive decline in orientation.**

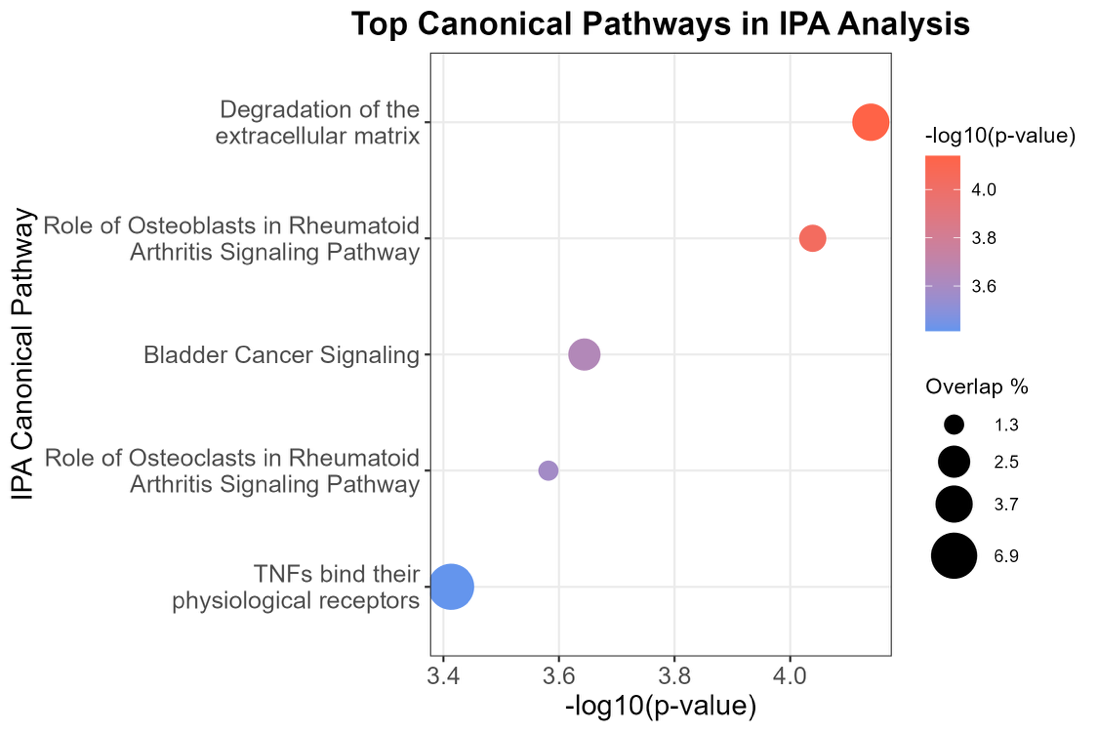

Top 5 canonical pathways identified by IPA. Dot represents overlap percentage, with colour representing statistical significance (−log₁₀ p-value) scaling from blue (less significant) to red (more significant). Statistically significant proteins (FDR < 0.05) associated with decline in orientation were used as input. Default settings used.

**Supplementary Figure 8. Top 5 upstream regulators identified by IPA associated with cognitive decline in orientation.**

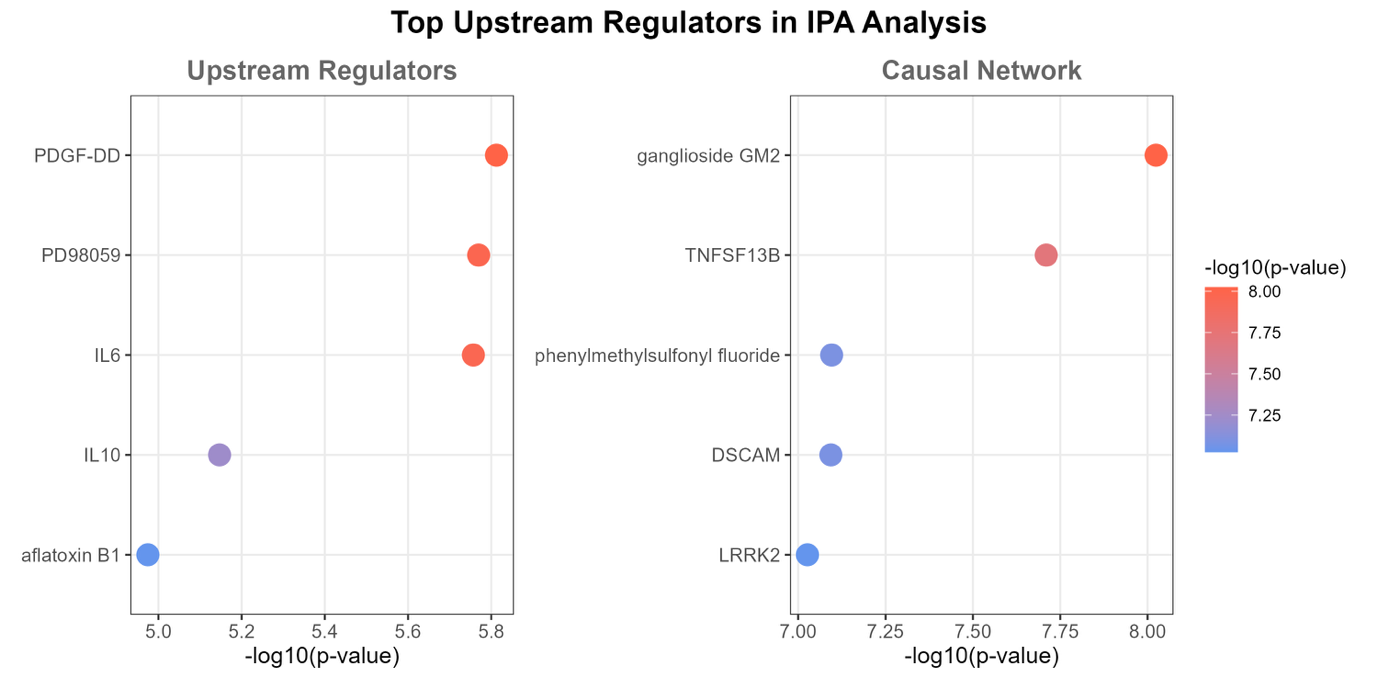

Top 5 upstream regulators identified by Ingenuity Pathway Analysis (IPA) for Upstream Regulators (left) and Causal Network (right). Dot represents statistical significance (−log₁₀ p-value), with colour scaling from blue (less significant) to red (more significant). Statistically significant proteins (FDR < 0.05) associated with decline in orientation were used as input. Default settings used.

**Supplementary Figure 9. Cell type expression from ROSMAP single-cell RNA-seq.**

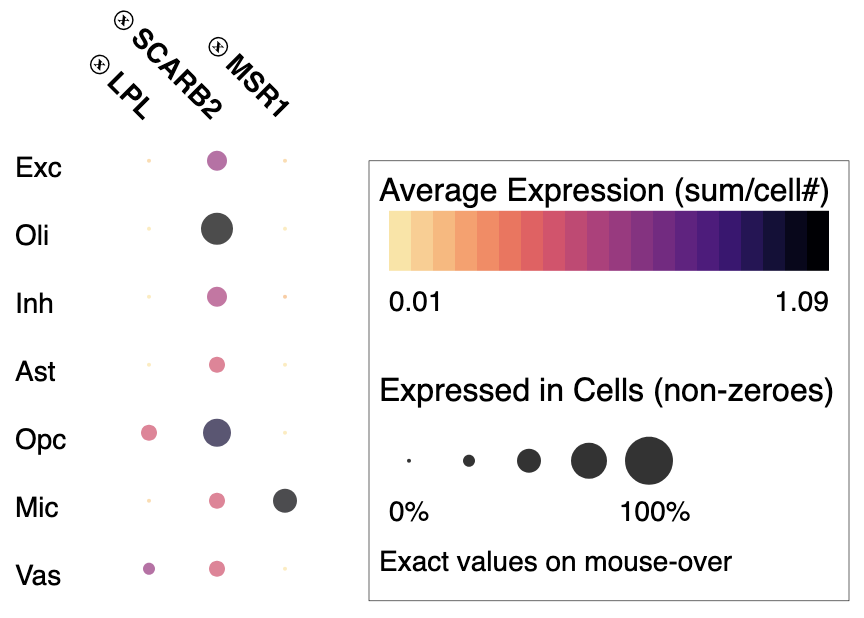

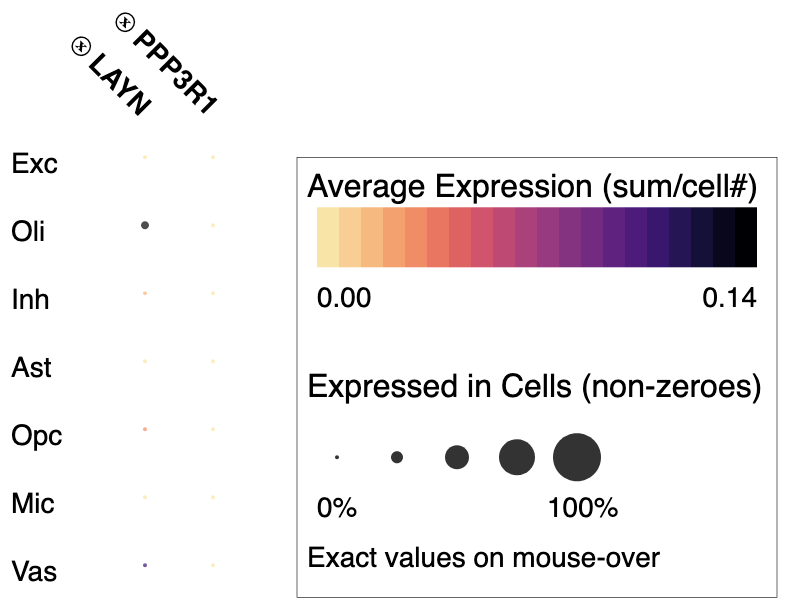

Genes encoding the proteins associated with decline in orientation are expressed by microglia, oligodendrocytes and the vasculature. Dot plot of expression with respect to the different cell-types identified in single-cell RNA-seq data of human aged and Alzheimer’s disease prefrontal cortexes from 92 individuals.[2, 3] *PPP3R1* is expressed at low levels by all cell types identified.

**Supplementary Figure 10. Cell type eQTLs.**

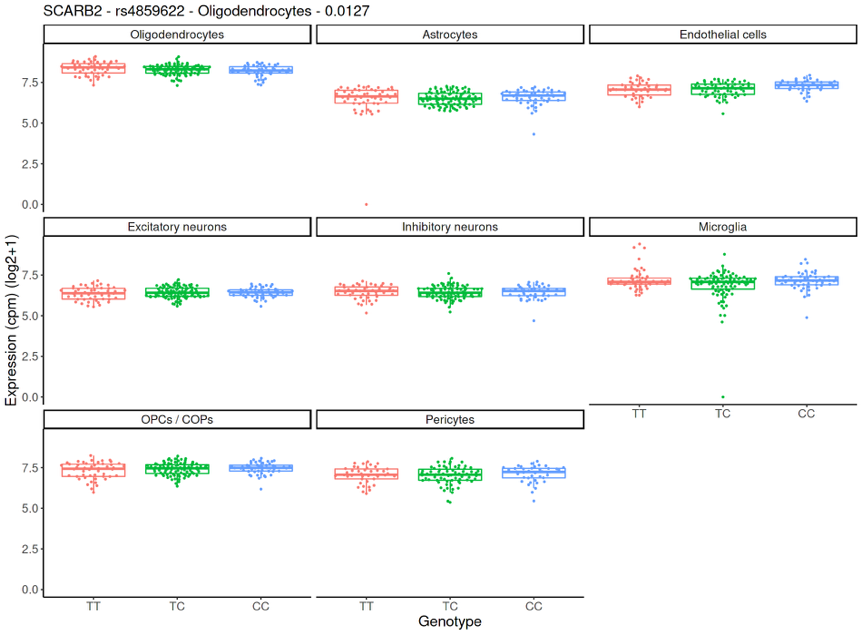

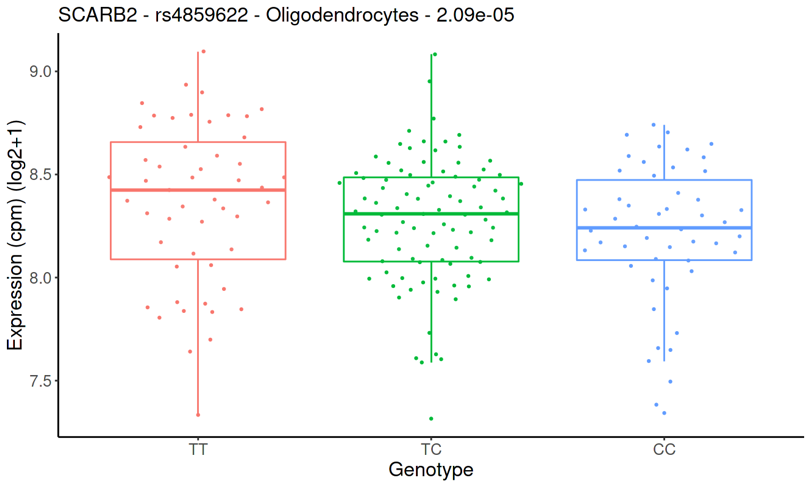

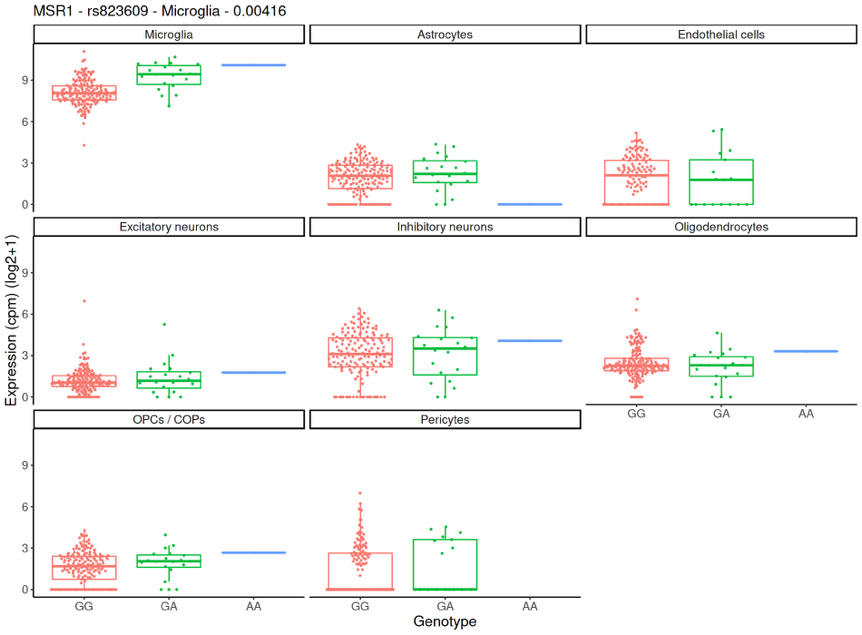

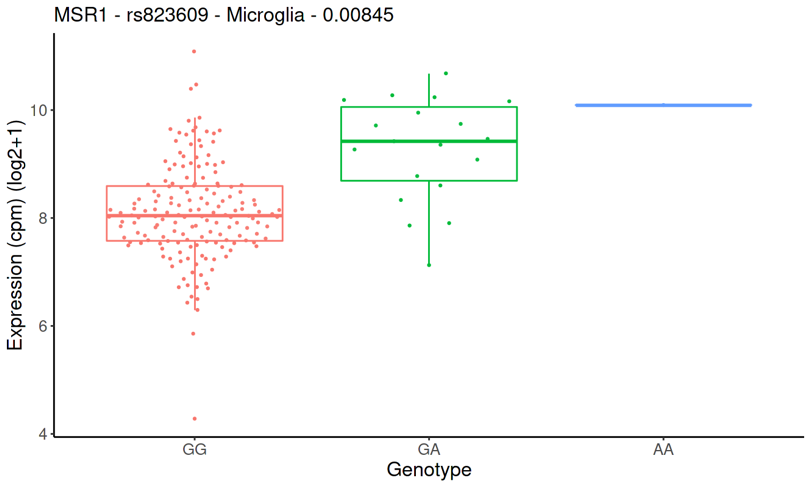

(Left) Cell-type-specific cis-eQTL for *SCARB2* and *MSR1* in oligodendrocytes and microglia respectively (n > 187 biologically independent individuals). Each dot represents an individual. The number top-left is a P-value (Wald z-statistic) testing whether the genetic effect in the discovered cell type (top-left cell type) is different from the genetic effects in all other cell types, and adjusted for multiple testing with the Benjamini–Hochberg procedure. (Right) Top cis-eQTLs for *SCARB2* and *MSR1* in oligodendrocytes and microglia (n >187 biologically independent individuals, each dot shown). The number above graph is a two-sided P-value obtained using fastQTL and corrected for multiple testing using the Benjamini–Hochberg procedure.[4]

**References**

[1] Kolberg, L., Raudvere, U., Kuzmin, I., Vilo, J., & Peterson, H. (2020). gprofiler2--an R package for gene list functional enrichment analysis and namespace conversion toolset g: Profiler. *F1000Research*, *9*, ELIXIR-709.

[2] Xiong, X., James, B. T., Boix, C. A., Park, Y. P., Galani, K., Victor, M. B., ... & Kellis, M. (2023). Epigenomic dissection of Alzheimer’s disease pinpoints causal variants and reveals epigenome erosion. *Cell*, *186*(20), 4422-4437.

[3] Mathys, H., Peng, Z., Boix, C. A., Victor, M. B., Leary, N., Babu, S., ... & Tsai, L. H. (2023). Single-cell atlas reveals correlates of high cognitive function, dementia, and resilience to Alzheimer’s disease pathology. *Cell*, *186*(20), 4365-4385.

[4] Bryois, J., Calini, D., Macnair, W., Foo, L., Urich, E., Ortmann, W., ... & Malhotra, D. (2022). Cell-type-specific cis-eQTLs in eight human brain cell types identify novel risk genes for psychiatric and neurological disorders. *Nature neuroscience*, *25*(8), 1104-1112.
